# A cross-tissue, age-specific flow cytometry reference for immune cells in the airways and blood of children

**DOI:** 10.1101/2024.05.08.24307019

**Authors:** Shivanthan Shanthikumar, Liam Gubbels, Karen Davies, Hannah Walker, Anson Tsz Chun Wong, Jovana Maksimovic, Alicia Oshlack, Richard Saffery, Eric Levi, Sarath C. Ranganathan, Melanie R. Neeland

## Abstract

Respiratory diseases are a common cause of morbidity and hospitalisation for children. Despite this, treatment options are limited and are often ineffective. The development of curative or disease-modifying treatments for children relies on a better understanding of underlying immunity in the early airway. To establish a flow cytometry reference for immune cells in the paediatric airway, we analysed 178 samples from 66 children aged between 1-15 years. This included five tissues of the upper (nasal brushings, palatine tonsils, adenotonsil) and lower (bronchial brushings, bronchoalveolar lavage (BAL)) airway, as well as whole blood for paired analysis of local and systemic immune response. Nasal, bronchial, and alveolar samples were analysed using a 17-plex antibody panel that captures cells of immune and epithelial lineage, while tonsil, adenoid, and blood samples were analysed using a 31-plex antibody panel that extensively phenotypes mononuclear immune cells. All protocols, panels, and data are openly available, to facilitate implementation in paediatric clinical laboratories. We provide age-specific cell reference data for infancy (0-2 years), preschool (3-5 years), childhood (6-10 years) and adolescence (11-15 years) for 37 cell populations. We show tissue-specific maturation of the airway immune system across childhood, further highlighting the importance of developing age-specific references of the paediatric airway. Intra-individual, cross-tissue analysis of paired samples revealed marked correlation in immune cell proportions between paired nasal-bronchial samples, paired tonsil-adenoid samples, and paired adenoid-blood samples, which may have implications for clinical testing. Our study advances knowledge of airway immunity from infancy through to adolescence and provides an openly available control dataset to aid in interpretation of clinical findings in samples obtained from children with respiratory diseases.

## INTRODUCTION

Respiratory diseases are a major source of morbidity and hospitalisation in children worldwide. They not only cause significant short-term effects but may also have lifelong impacts on lung function ^1^. Despite this, treatment options for children are limited, largely extrapolated from adults, and often confer little to no clinical benefit. This is due, in part, to limited understanding of the mechanisms that underly disease in children and a lack of age- specific reference ranges for cells and soluble factors present in the paediatric airway.

A lack of age-specific, reliable reference ranges for children extends beyond the respiratory system into all aspects of paediatric health care ^2, 3^. Many of the available reference intervals were determined over three decades ago using technologies that are no longer relevant in a clinical setting. Despite considerable efforts to address this in recent years, current paediatric reference ranges remain largely incomplete ^4^. The development of age-specific reference ranges is particularly crucial in paediatric respiratory medicine due to the marked age-related differences in susceptibility and immunological response to a range of infectious and chronic conditions of the lung. This was recently highlighted during the COVID-19 pandemic, with children showing a markedly mild manifestation of disease compared to adults ^5^, and is also observed for other conditions including RSV infection ^6^, wheezing, and asthma ^7^. A better understanding of immune cell composition in the healthy paediatric airway may help to identify diagnostic biomarkers of disease, targets for anti-inflammatory therapy, and inform novel interventions such as mucosal vaccination.

We aimed to develop a clinically relevant age-specific flow cytometry reference for immune cells in the upper and lower airways of children. Our secondary aims were to explore changes in immune cell composition in the first 15 years of life, understand the relationship between airway and systemic immunity, and discover cross-tissue relationships in immune cell proportions within individuals.

## RESULTS

### 1. Establishing a clinical reference for immune cells in airways and blood of children

To establish a reference for immune cells in the paediatric airway, we sampled five tissues comprising both the upper airway (nasal brushings, palatine tonsils, adenotonsil), and lower airway (bronchial brushings, BAL) from 66 children undergoing general anaesthesia at the Royal Children’s Hospital Melbourne. The clinical indications for these procedures are provided in Supplementary Table 1, with >80% of the cohort undergoing a procedure for obstructive sleep apnoea (OSA). Medical history and information on respiratory status from parent questionnaires are provided in Supplementary Table 2. These children were aged between 1 and 15 years, allowing for age stratification across infancy (0-2 years), preschool period (3-5 years), childhood (6-10 years), and adolescence (11-15 years) (Figure 1A). Furthermore, where possible, multiple airway samples were collected per participant, allowing for intra-individual, cross-tissue assessment of immunity in the upper and lower airway. We also analysed whole blood in order to establish a reference panel of circulating immune cells and to enable associations between the systemic and respiratory immune systems in childhood. Airway and blood samples were processed to single-cell suspensions and analysed by spectral flow cytometry using two multiplexed panels (Figure 1B). The first panel, comprising 17 markers, captures cell populations present in nasal, bronchial, and alveolar samples (Supplementary Table 3). These cell types include those of both immune and epithelial lineage (Figure 1C). The second panel, comprising 31 markers, captures immune cell populations present in the palatine tonsil (“tonsil”), adenotonsil (“adenoid”), and blood samples (Supplementary Table 4). These include all major immune cell populations and their subtypes, with a focus on B- and T- cell lineages given the role of tonsils and adenoids as major lymphoid niches (Figure 1D). We designed publicly available sample processing and flow cytometry protocols (protocols.io/workspaces/earlyAIR) with the goal they could be implemented in paediatric clinical laboratories.

**Figure 1.**
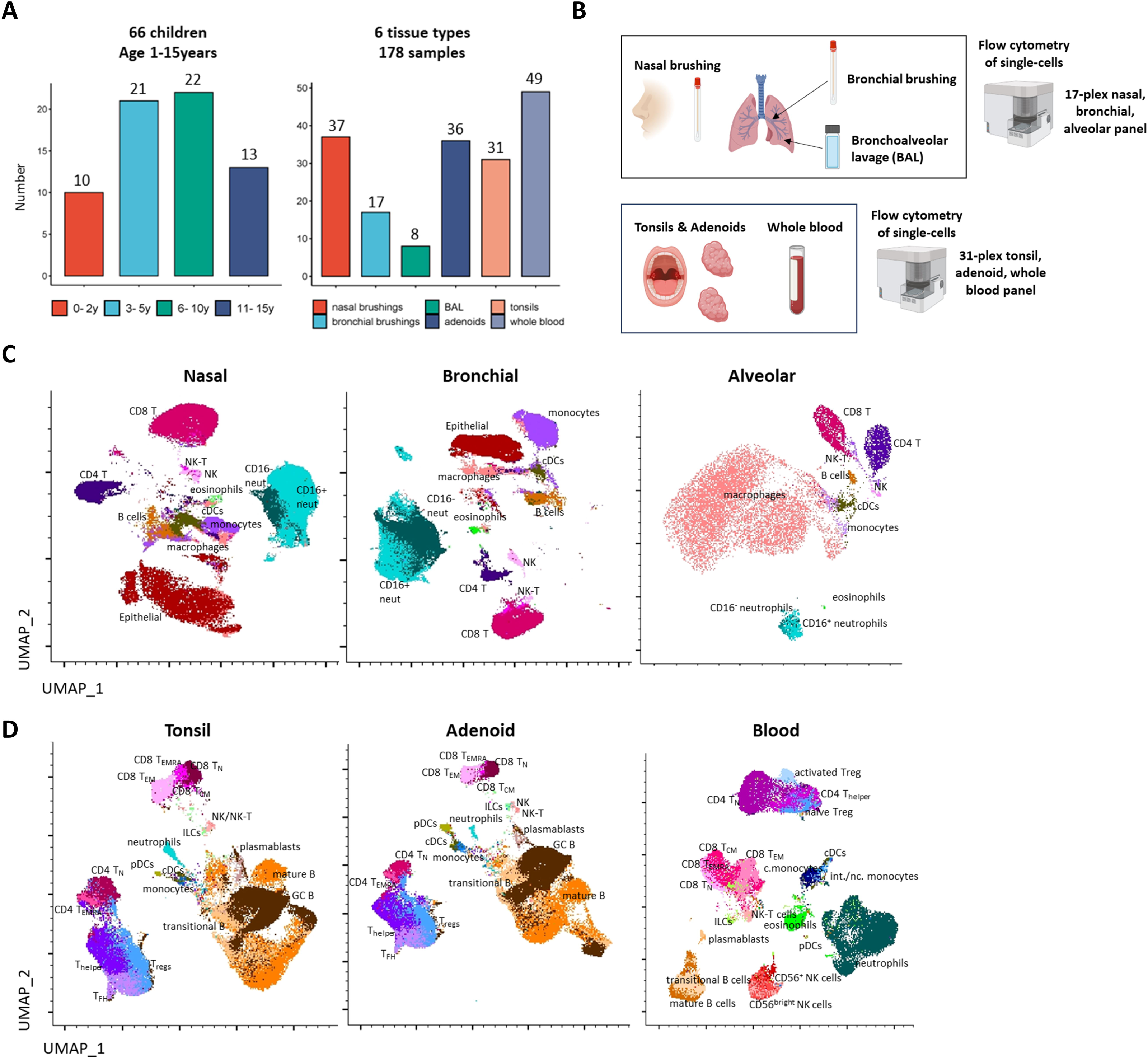
Overview of the experimental design and cellular landscape of each tissue. **(A)** 66 children aged between 1 and 15 years were recruited into the earlyAIR study. There were 10 infants (age 0-2 years), 21 preschoolers (age 3-5 years), 22 children (6-10 years), and 13 adolescents (11-15 years). From these 66 children, 178 samples were collected across six tissue types: nasal brushings (n=37), bronchial brushings (n=17), bronchoalveolar lavage (BAL) (n=8), adenoids (n=36), tonsils (n=31), and whole blood (n=49). **(B)** Each sample type was processed into a single-cell suspension and analysed by spectral flow cytometry. Nasal brushing, bronchial brushing, and BAL samples were analysed using a 17-plex lung panel. Tonsil, adenoid, and whole blood samples were analysed using a 31-plex tonsil-adenoid-blood panel. **(C)** Uniform Manifold Approximation and Projection (UMAP) plots showing the cellular landscape of nasal, bronchial and alveolar cells identified using the lung panel. **(D)** UMAP plots showing the cellular landscape of tonsil, adenoid, and blood cells identified using the tonsil-adenoid-blood panel.

### 2. Immune cell landscape of paediatric nasal, bronchial, and alveolar samples

Epithelial and immune cell populations were identified in nasal, bronchial, and alveolar samples based on expression of 16 key lineage markers (Figure 2A-C, Supplementary Figure 1). Immune cell populations identified were neutrophils, eosinophils, macrophages, NK cells, NK-T cells, CD8 T cells, CD4 T cells, B cells, monocytes, and conventional dendritic cells (cDCs). In all compartments, the immune cell lineage outnumbered the epithelial lineage, particularly more distally through the airway (Figure 2D-F). In nasal samples, immune cells comprised a median of 74.3% of all cells, bronchial samples 88%, and alveolar samples 98% (Figure 2D-F). Median proportions of the 10 major immune cell subtypes in each sample type are shown in Figure 2G-I. To help develop age-specific references for each of these tissues, the median and range of each cell population are reported by clinically relevant age groups (all ages (age 1-15 years), infancy (0-2 years), preschool (3-5 years), childhood (6-10 years), adolescence (11-15 years)) and presented in Tables 1-3. For nasal samples, all five age analyses are reported (Table 1). For bronchial and alveolar samples, due to more limited numbers, a subset of these age groups is reported (Tables 2-3).

**Figure 2.**
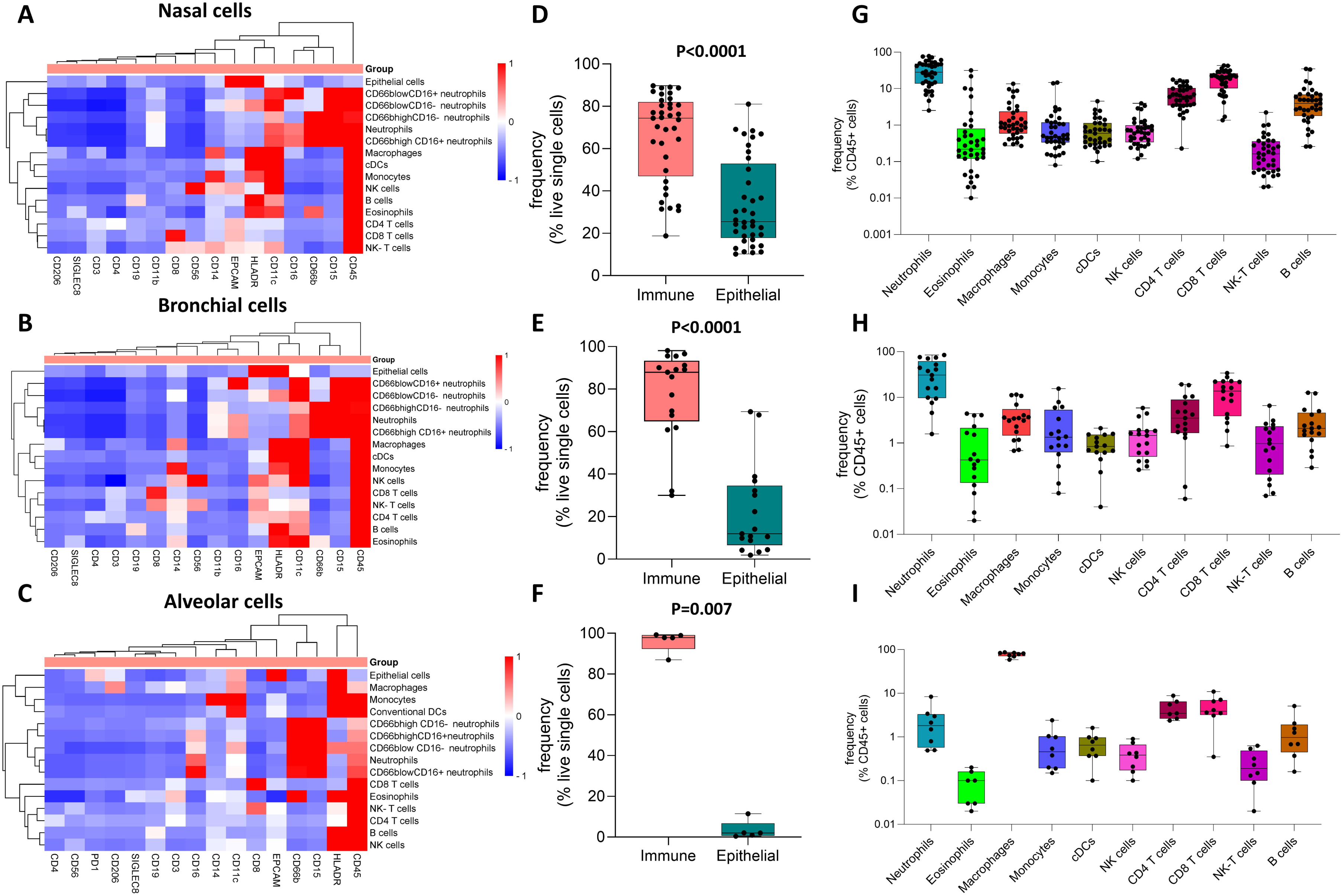
Marker expression and proportion of cell populations identified in paediatric nasal, bronchial, and alveolar samples. **(A)** Heatmap depicting hierarchical clustering analysis of the normalised median fluorescence intensity (MFI) of 16 lineage markers on cell types identified in nasal brushings samples. **(B)** Heatmap depicting hierarchical clustering analysis of the normalised MFI of 16 lineage markers on cell types identified in bronchial brushings samples. **(C)** Heatmap depicting hierarchical clustering analysis of the normalised MFI of 16 lineage markers on cell types identified in BAL samples. **(D)** Proportion of immune and epithelial linages in nasal brushings samples. **(E)** Proportion of immune and epithelial linages in bronchial brushings samples. **(F)** Proportion of immune and epithelial linages in BAL samples. **(G)** Box plots depicting individual cell proportions for 10 broad immune cell types identified in nasal brushings samples **(H)** Box plots depicting individual cell proportions for 10 broad immune cell types identified in bronchial brushings samples. **(I)** Box plots depicting individual cell proportions for 10 broad immune cell types identified in BAL samples. P- values determined by Mann-Whitney U-test where p<0.05 was considered significant.

**Table 1.** Immune cell subtype frequency in nasal brushings samples from children aged 0-15 years.

| **NASAL**<br>**IMMUNE CELLS** | **0-15 years**<br>**(n=37)** | **0-2 years**<br>**(n=5)** | **3-5 years**<br>**(n=16)** | **6-10 years**<br>**(n=12)** | **11-15 years**<br>**(n=4)** |
| --- | --- | --- | --- | --- | --- |
| **Total granulocytes**<br>%CD45<sup>+</sup> cells<br>Median (range) | 31.72<br>(2.92-88.20) | 29.09<br>(23.42-79.66) | 46.03<br>(6.93-79.63) | 20.40<br>(2.92-88.20) | 51.67<br>(42.28-64.37) |
| **Eosinophils**<br>%CD45<sup>+</sup> cells<br>Median (range) | 0.22<br>(0.01-31.62) | 0.34<br>(0.16-0.50) | 0.16<br>(0.02-2.16) | 0.61<br>(0.01-31.62) | 0.63<br>(0.02-21.92) |
| **Neutrophils**<br>%CD45<sup>+</sup> cells<br>Median (range) | 28.17<br>(2.51-78.26) | 28.17<br>(20.55-71.76) | 42.84<br>(6.83-78.26) | 14.35<br>(2.51-60.89) | 43.52<br>(39.82-52.74) |
| **Macrophages**<br>%CD45<sup>+</sup> cells<br>Median (range) | 0.99<br>(0.27-13.63) | 1.16<br>(0.30-8.37) | 0.78<br>(0.27-9.69) | 1.14<br>(0.46-13.63) | 1.16<br>(0.79-3.29) |
| **NK cells**<br>%CD45<sup>+</sup> cells<br>Median (range) | 0.64<br>(0.12-3.96) | 1.03<br>(0.48-2.9) | 0.74<br>(0.12-3.96) | 0.42<br>(0.15-3.39) | 0.59<br>(0.31-0.60) |
| **NK-T cells**<br>%CD45<sup>+</sup> cells<br>Median (range) | 0.16<br>(0.01-2.23) | 0.27<br>(0.06-2.23) | 0.10<br>(0.04-1.02) | 0.13<br>(0.01-0.62) | 0.18<br>(0.02-0.23) |
| **CD8 T cells**<br>%CD45<sup>+</sup> cells<br>Median (range) | 20.71<br>(1.36-43.14) | 20.25<br>(6.48-31.47) | 19.09<br>(1.36-43.14) | 22.12<br>(1.71-31.69) | 24.29<br>(13.34-35.89) |
| **CD4 T cells**<br>%CD45<sup>+</sup> cells<br>Median (range) | 5.91<br>(0.23-17.72) | 8.94<br>(0.23-17.72) | 5.19<br>(1.54-14.64) | 9.33<br>(1.91-16.90) | 4.32<br>(3.46-7.00) |
| **B cells**<br>%CD45<sup>+</sup> cells<br>Median (range) | 3.92<br>(0.26-35.02) | 2.82<br>(0.26-7.07) | 5.51<br>(2.57-35.02) | 3.73<br>(0.54-33.71) | 1.53<br>(0.26-2.64) |
| **Monocytes**<br>%CD45<sup>+</sup> cells<br>Median (range) | 0.50<br>(0.08-14.72) | 1.20<br>(0.08-14.06) | 0.48<br>(0.14-3.08) | 0.46<br>(0.15-14.72) | 0.33<br>(0.16-2.72) |
| **cDCs**<br>%CD45<sup>+</sup> cells<br>Median (range) | 0.47<br>(0.01-4.55) | 1.57<br>(0.01-4.55) | 0.43<br>(0.01-2.66) | 0.36<br>(0.23-2.07) | 0.63<br>(0.43-1.14) |

**Table 2.** Immune cell subtype frequency in bronchial brushings samples from children aged 0-15 years.

| **BRONCHIAL**<br>**IMMUNE CELLS** | **0-15 years**<br>**(n=17)** | **0-5 years**<br>**(n=8)** | **6-10 years**<br>**(n=5)** | **11-15 years**<br>**(n=4)** |
| --- | --- | --- | --- | --- |
| **Total granulocytes**<br>%CD45<sup>+</sup> cells<br>Median (range) | 40.84<br>(1.85-89.57) | 58.86<br>(5.04-89.57) | 18.34<br>(1.85-56.76) | 33.53<br>(15.00-47.55) |
| **Eosinophils**<br>%CD45<sup>+</sup> cells<br>Median (range) | 0.41<br>(0.01-4.41) | 0.23<br>(0.02-1.52) | 1.66<br>(0.12-4.04) | 2.34<br>(0.01-4.41) |
| **Neutrophils**<br>%CD45<sup>+</sup> cells<br>Median (range) | 30.75<br>(1.58-85.04) | 56.10<br>(4.6-85.04) | 15.91<br>(1.58-52.41) | 20.41<br>(9.88-37.80) |
| **Macrophages**<br>%CD45<sup>+</sup> cells<br>Median (range) | 3.46<br>(0.68-11.56) | 1.47<br>(0.68-3.71) | 3.53<br>(3.09-10.74) | 9.03<br>(3.62-11.56) |
| **NK cells**<br>%CD45<sup>+</sup> cells<br>Median (range) | 1.46<br>(0.26-5.88) | 1.24<br>(0.31-5.88) | 0.98<br>(0.26-1.83) | 2.74<br>(0.61-4.44) |
| **NK-T cells**<br>%CD45<sup>+</sup> cells<br>Median (range) | 0.98<br>(0.07-6.56) | 0.33<br>(0.07-2.05) | 1.70<br>(0.16-2.65) | 2.60<br>(0.78-6.56) |
| **CD8 T cells**<br>%CD45<sup>+</sup> cells<br>Median (range) | 13.78<br>(0.86-33.92) | 7.89<br>(1.85-21.82) | 13.78<br>(0.86-33.92) | 22.56<br>(8.43-27.76) |
| **CD4 T cells**<br>%CD45<sup>+</sup> cells<br>Median (range) | 3.35<br>(0.06-19.24) | 3.77<br>(0.11-19.24) | 4.23<br>(1.58-10.50) | 6.49<br>(0.06-18.61) |
| **B cells**<br>%CD45<sup>+</sup> cells<br>Median (range) | 2.10<br>(0.29-12.61) | 2.54<br>(0.5-5.35) | 1.86<br>(0.29-12.26) | 1.87<br>(0.67-12.61) |
| **Monocytes**<br>%CD45<sup>+</sup> cells<br>Median (range) | 1.27<br>(0.01-15.58) | 1.53<br>(0.08-15.58) | 1.59<br>(0.31-5.95) | 0.73<br>(0.01-1.39) |
| **cDCs**<br>%CD45<sup>+</sup> cells<br>Median (range) | 0.70<br>(0.01-2.11) | 0.73<br>(0.01-2.11) | 0.70<br>(0.04-1.59) | 0.87<br>(0.01-1.52) |

**Table 3.**
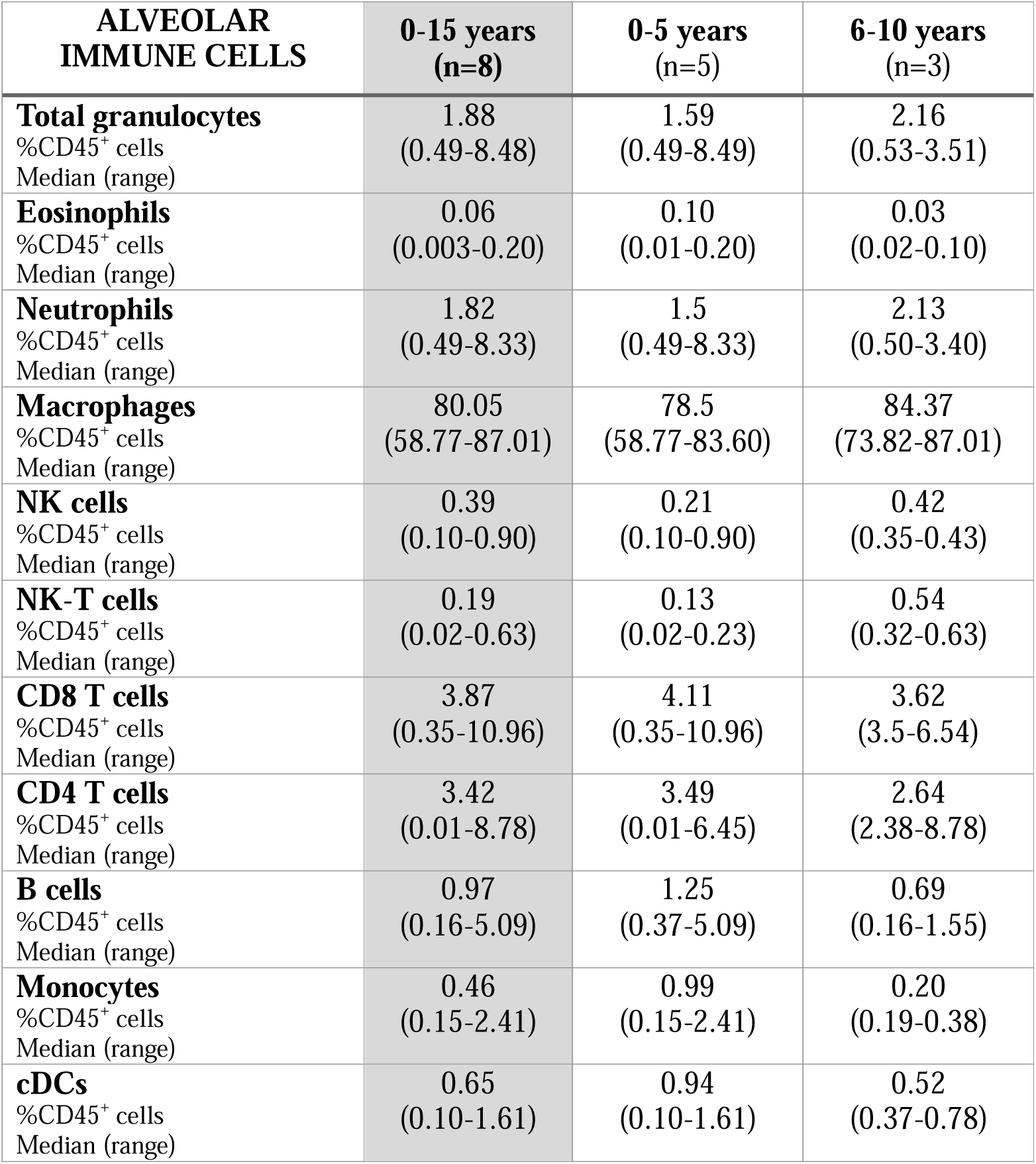
Immune cell subtype frequency in bronchoalveolar lavage (BAL) samples from children aged 0-15 years.

| **ALVEOLAR**<br>**IMMUNE CELLS** | **0-15 years**<br>**(n=8)** | **0-5 years**<br>**(n=5)** | **6-10 years**<br>**(n=3)** |
| --- | --- | --- | --- |
| **Total granulocytes**<br>%CD45<sup>+</sup> cells<br>Median (range) | 1.88<br>(0.49-8.48) | 1.59<br>(0.49-8.49) | 2.16<br>(0.53-3.51) |
| **Eosinophils**<br>%CD45<sup>+</sup> cells<br>Median (range) | 0.06<br>(0.003-0.20) | 0.10<br>(0.01-0.20) | 0.03<br>(0.02-0.10) |
| **Neutrophils**<br>%CD45<sup>+</sup> cells<br>Median (range) | 1.82<br>(0.49-8.33) | 1.5<br>(0.49-8.33) | 2.13<br>(0.50-3.40) |
| **Macrophages**<br>%CD45<sup>+</sup> cells<br>Median (range) | 80.05<br>(58.77-87.01) | 78.5<br>(58.77-83.60) | 84.37<br>(73.82-87.01) |
| **NK cells**<br>%CD45<sup>+</sup> cells<br>Median (range) | 0.39<br>(0.10-0.90) | 0.21<br>(0.10-0.90) | 0.42<br>(0.35-0.43) |
| **NK-T cells**<br>%CD45<sup>+</sup> cells<br>Median (range) | 0.19<br>(0.02-0.63) | 0.13<br>(0.02-0.23) | 0.54<br>(0.32-0.63) |
| **CD8 T cells**<br>%CD45<sup>+</sup> cells<br>Median (range) | 3.87<br>(0.35-10.96) | 4.11<br>(0.35-10.96) | 3.62<br>(3.5-6.54) |
| **CD4 T cells**<br>%CD45<sup>+</sup> cells<br>Median (range) | 3.42<br>(0.01-8.78) | 3.49<br>(0.01-6.45) | 2.64<br>(2.38-8.78) |
| **B cells**<br>%CD45<sup>+</sup> cells<br>Median (range) | 0.97<br>(0.16-5.09) | 1.25<br>(0.37-5.09) | 0.69<br>(0.16-1.55) |
| **Monocytes**<br>%CD45<sup>+</sup> cells<br>Median (range) | 0.46<br>(0.15-2.41) | 0.99<br>(0.15-2.41) | 0.20<br>(0.19-0.38) |
| **cDCs**<br>%CD45<sup>+</sup> cells<br>Median (range) | 0.65<br>(0.10-1.61) | 0.94<br>(0.10-1.61) | 0.52<br>(0.37-0.78) |

### 3. Immune cell landscape of paediatric tonsil, adenoid, and blood samples

Immune cell populations were identified in tonsil, adenoid and whole blood samples based on expression of 22 lineage markers (Figure 3A-C, Supplementary Figure 2). In tonsils and adenoids, 25 immune cell populations were quantified, and in whole blood 30 immune cell populations were quantified. Additional populations observed in blood that were not observed in tonsil/adenoid tissue include eosinophils, CD56^bright^ subsets of NK cells and subsets of CD16 expressing monocytes including non-classical and intermediate monocytes. On the other hand, germinal centre (GC) B cells, one of the most abundant populations in tonsils and adenoids, were not observed in blood. Mean Fluorescence Intensity (MFI) analysis of marker expression revealed differential surface receptor patterns across the three compartments including: CD24 expression on NK and NK-T cells from adenoid and tonsil tissue that is not observed on the respective blood populations; high expression of CD38 on T cell populations in tissue: and CXCR5 expression NK-T cells in tissue (Figure 3A-C). Median proportions of immune cell populations in all tonsil, adenoid, and blood samples are shown in Figure 3D-F, respectively. To help develop age-specific references for each of these tissues, the median and range of each cell population are reported by clinically relevant age groups (all ages, infancy, preschool, childhood, adolescence) and presented in Tables 4-6.

**Figure 3.**
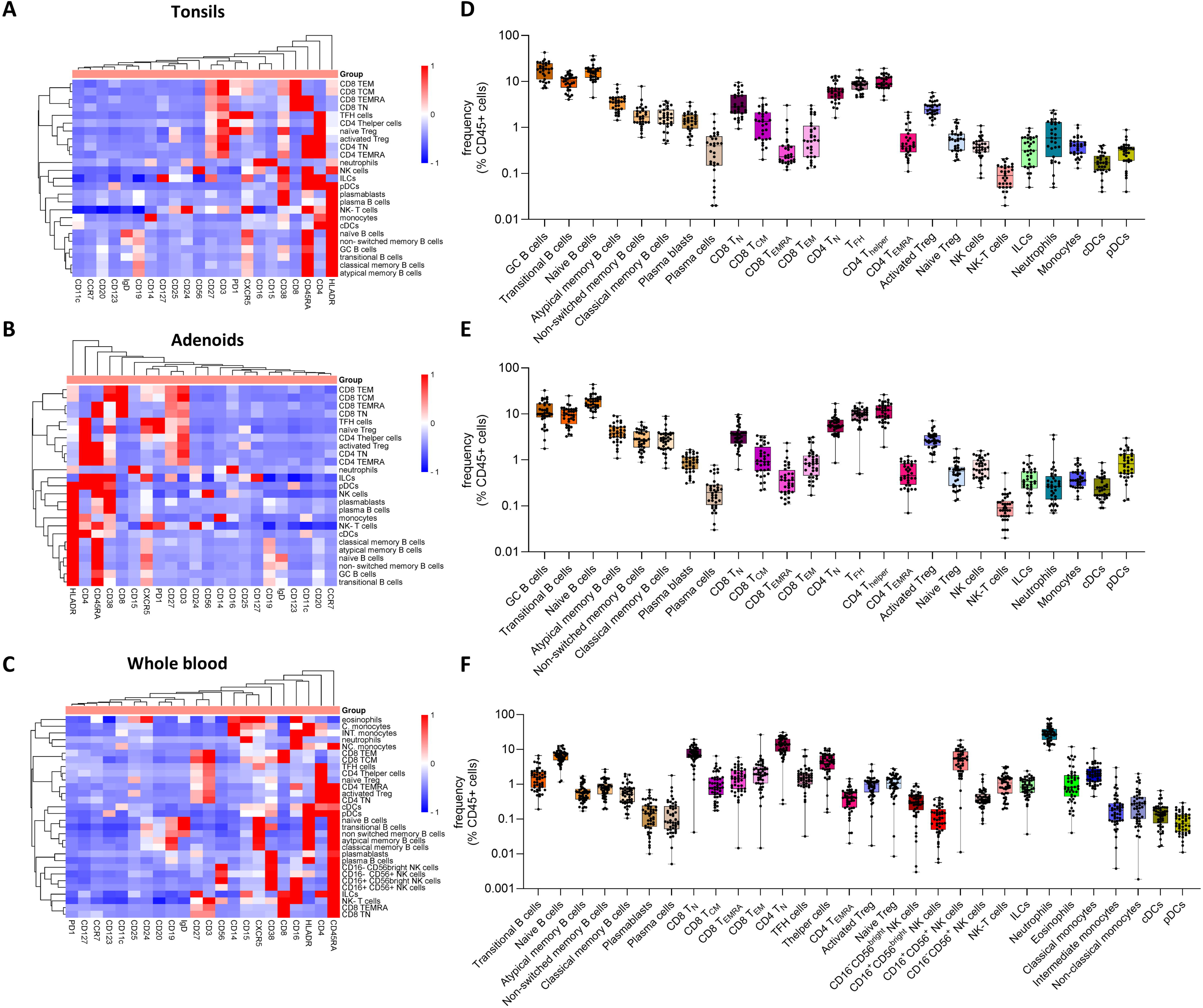
Marker expression and proportion of cell populations identified in paediatric tonsil, adenoid, and whole blood samples. **(A)** Heatmap depicting hierarchical clustering analysis of the normalised median fluorescence intensity (MFI) of 22 lineage markers on cell types identified in tonsils. **(B)** Heatmap depicting hierarchical clustering analysis of the normalised MFI of 22 lineage markers on cell types identified in adenoids. **(C)** Heatmap depicting hierarchical clustering analysis of the normalised MFI of 22 lineage markers on cell types identified in blood. **(D)** Box plots depicting individual cell proportions for 25 immune cell types identified in tonsils. **(E)** Box plots depicting individual cell proportions for 25 immune cell types identified in adenoids. **(F)** Box plots depicting individual cell proportions for 30 immune cell types identified in whole blood samples.

**Table 4.** Immune cell subtype frequency in tonsils from children aged 0-15 years.

| **TONSIL**<br>**IMMUNE CELLS** | **0-15 years**<br>**(n=31)** | **0-2 years**<br>**(n=5)** | **3-5 years**<br>**(n=13)** | **6-10 years**<br>**(n=9)** | **11-15 years**<br>**(n=4)** |
| --- | --- | --- | --- | --- | --- |
| **Neutrophils**<br>%CD45<sup>+</sup> cells<br>Median (range) | 0.59<br>(0.05-2.36) | 0.17<br>(0.06-1.29) | 0.70<br>(0.05-1.52) | 0.49<br>(0.19-1.18) | 2.03<br>(1.34-2.36) |
| **Plasma blasts**<br>%CD45<sup>+</sup> cells<br>Median (range) | 1.38<br>(0.41-3.54) | 1.53<br>(1.20-1.95) | 1.33<br>(0.54-2.91) | 1.60<br>(0.41-3.54) | 1.32<br>(0.94-1.81) |
| **GC B cells**<br>%CD45<sup>+</sup> cells<br>Median (range) | 18.84<br>(7.16-42.73) | 22.69<br>(7.16-42.73) | 18.47<br>(9.64-30.00) | 19.01<br>(9.31-27.40) | 9.59<br>(7.25-24.30) |
| **Atypical memory B cells**<br>%CD45<sup>+</sup> cells<br>Median (range) | 3.62<br>(1.44-8.64) | 2.19<br>(1.78-2.98) | 4.42<br>(2.47-7.01) | 3.62<br>(1.82-4.50) | 2.79<br>(1.44-8.64) |
| **Classical memory B cells**<br>%CD45<sup>+</sup> cells<br>Median (range) | 1.63<br>(0.45-3.72) | 0.53<br>(0.45-1.60) | 1.80<br>(0.79-2.59) | 1.47<br>(0.77-3.72) | 2.77<br>(1.56-3.39) |
| **Naïve B cells**<br>%CD45<sup>+</sup> cells<br>Median (range) | 16.45<br>(4.44-36.15) | 14.39<br>(9.73-29.40) | 15.07<br>(9.93-19.94) | 16.53<br>(4.44-29.87) | 18.44<br>(17.06-36.15) |
| **Non-switched memory B cells**<br>%CD45<sup>+</sup> cells<br>Median (range) | 1.69<br>(0.61-7.88) | 1.29<br>(0.74-1.92) | 1.77<br>(0.87-3.99) | 1.69<br>(0.61-7.88) | 2.51<br>(1.30-2.77) |
| **Plasma cells**<br>%CD45<sup>+</sup> cells<br>Median (range) | 0.40<br>(0.02-1.96) | 0.16<br>(0.03-1.31) | 0.40<br>(0.04-1.96) | 0.45<br>(0.15-0.64) | 0.10<br>(0.02-0.70) |
| **Transitional B cells**<br>%CD45<sup>+</sup> cells<br>Median (range) | 10.27<br>(4.06-17.21) | 14.81<br>(6.50-17.21) | 11.33<br>(7.36-14.24) | 8.76<br>(4.06-13.25) | 5.90<br>(5.04-10.02) |
| **NK cells**<br>%CD45<sup>+</sup> cells<br>Median (range) | 0.37<br>(0.08-1.09) | 0.37<br>(0.22-0.80) | 0.37<br>(0.1-1.09) | 0.37<br>(0.08-0.59) | 0.37<br>(0.24-0.50) |
| **NK-T cells**<br>%CD45<sup>+</sup> cells<br>Median (range) | 0.09<br>(0.02-0.21) | 0.07<br>(0.02-0.21) | 0.09<br>(0.04-0.20) | 0.11<br>(0.03-0.18) | 0.05<br>(0.04-0.12) |
| **CD8 TCM**<br>%CD45<sup>+</sup> cells<br>Median (range) | 1.35<br>(0.2-4.34) | 0.41<br>(0.20-1.38) | 1.11<br>(0.33-4.34) | 1.35<br>(0.46-3.11) | 2.38<br>(1.94-2.81) |
| **CD8 TN**<br>%CD45<sup>+</sup> cells<br>Median (range) | 2.79<br>(0.94-9.54) | 2.83<br>(1.6-9.54) | 3.21<br>(1.51-7.71) | 2.97<br>(1.43-8.29) | 1.73<br>(0.94-2.79) |
| **CD8 TEMRA**<br>%CD45<sup>+</sup> cells<br>Median (range) | 0.23<br>(0.12-3.04) | 0.22<br>(0.17-0.38) | 0.26<br>(0.14-0.93) | 0.20<br>(0.36-2.17) | 0.19<br>(0.12-0.41) |
| **CD8 TEM**<br>%CD45<sup>+</sup> cells<br>Median (range) | 0.52<br>(0.13-3.01) | 0.23<br>(0.14-0.33) | 0.64<br>(0.13-2.13) | 0.53<br>(0.36-2.17) | 1.66<br>(0.52-3.01) |
| **Activated Tregs**<br>%CD45<sup>+</sup> cells<br>Median (range) | 0.55<br>(0.19-1.47) | 0.41<br>(0.19-1.25) | 0.54<br>(0.20-1.47) | 0.57<br>(0.32-0.95) | 0.35<br>(0.31-0.57) |
| **Naive Tregs**<br>%CD45<sup>+</sup> cells<br>Median (range) | 2.46<br>(1.13-5.75) | 2.76<br>(1.95-3.10) | 2.46<br>(1.56-5.75) | 2.14<br>(1.43-4.78) | 3.01<br>(1.13-3.62) |
| **TFH cells**<br>%CD45<sup>+</sup> cells<br>Median (range) | 8.57<br>(4.59-17.84) | 8.49<br>(6.88-12.10) | 8.87<br>(4.66-11.77) | 8.56<br>(4.63-12.12) | 7.20<br>(4.59-17.84) |
| **CD4 Thelper cells**<br>%CD45<sup>+</sup> cells<br>Median (range) | 9.19<br>(3.93-19.32) | 7.60<br>(3.93-8.83) | 8.78<br>(7.16-14.70) | 12.10<br>(7.25-15.06) | 13.75<br>(11.69-19.32) |
| **CD4 TN**<br>%CD45<sup>+</sup> cells<br>Median (range) | 6.07<br>(1.61-13.14) | 5.65<br>(3.32-12.31) | 6.07<br>(2.60-10.50) | 6.65<br>(3.34-13.14) | 4.35<br>(1.61-6.59) |
| **CD4 TEMRA**<br>%CD45<sup>+</sup> cells<br>Median (range) | 0.39<br>(0.11-2.15) | 0.32<br>(0.21-0.89) | 0.44<br>(0.17-1.59) | 0.35<br>(0.19-2.15) | 0.79<br>(0.11-1.28) |
| **Monocytes**<br>%CD45<sup>+</sup> cells<br>Median (range) | 0.40<br>(0.13-1.13) | 0.22<br>(0.13-0.63) | 0.47<br>(0.26-1.13) | 0.28<br>(0.20-0.60) | 0.41<br>(0.28-0.51) |
| **cDCs**<br>%CD45<sup>+</sup> cells<br>Median (range) | 0.17<br>(0.04-0.41) | 0.16<br>(0.12-0.22) | 0.16<br>(0.10-0.35) | 0.18<br>(0.04-0.41) | 0.17<br>(0.12-0.39) |
| **pDCs**<br>%CD45<sup>+</sup> cells<br>Median (range) | 0.32<br>(0.04-0.89) | 0.26<br>(0.10-0.35) | 0.35<br>(0.19-0.89) | 0.28<br>(0.04-0.54) | 0.15<br>(0.09-0.30) |
| **ILCs**<br>%CD45<sup>+</sup> cells<br>Median (range) | 0.32<br>(0.05-0.96) | 0.39<br>(0.10-0.48) | 0.48<br>(0.08-0.96) | 0.31<br>(0.05-0.81) | 0.45<br>(0.27-0.60) |

**Table 5.** Immune cell subtype frequency in adenoids from children aged 0-15 years.

| **ADENOID**<br>**IMMUNE CELLS** | **0-15 years**<br>**(n=36)** | **0-2 years**<br>**(n=4)** | **3-5 years**<br>**(n=14)** | **6-10 years**<br>**(n=10)** | **11-15 years**<br>**(n=8)** |
| --- | --- | --- | --- | --- | --- |
| **Neutrophils**<br>%CD45<sup>+</sup> cells<br>Median (range) | 0.27<br>(0.07-3.47) | 0.48<br>(0.21-0.88) | 0.29<br>(0.11-2.83) | 0.26<br>(0.08-0.40) | 0.11<br>(0.07-3.47) |
| **Plasma blasts**<br>%CD45<sup>+</sup> cells<br>Median (range) | 0.89<br>(0.31-1.66) | 0.83<br>(0.55-1.30) | 1.02<br>(0.44-1.52) | 0.68<br>(0.31-1.46) | 0.90<br>(0.33-1.66) |
| **GC B cells**<br>%CD45<sup>+</sup> cells<br>Median (range) | 10.38<br>(1.76-32.58) | 11.33<br>(8.88-16.06) | 14.27<br>(9.15-32.58) | 9.41<br>(4.31-21.51) | 6.49<br>(1.76-19.93) |
| **Atypical memory B cells**<br>%CD45<sup>+</sup> cells<br>Median (range) | 3.85<br>(1.08-9.17) | 2.27<br>(1.73-4.15) | 4.59<br>(2.77-7.08) | 3.51<br>(2.46-6.92) | 3.87<br>(1.08-9.17) |
| **Classical memory B cells**<br>%CD45<sup>+</sup> cells<br>Median (range) | 2.80<br>(0.66-8.91) | 0.98<br>(0.66-1.32) | 2.88<br>(1.37-5.88) | 3.25<br>(1.82-7.87) | 3.14<br>(1.74-8.91) |
| **Naïve B cells**<br>%CD45<sup>+</sup> cells<br>Median (range) | 18.63<br>(8.30-43.75) | 22.71<br>(11.05-27.67) | 17.48<br>(11.78-36.03) | 17.32<br>(8.30-26.74) | 17.89<br>(11.17-43.75) |
| **Non-switched memory B cells**<br>%CD45<sup>+</sup> cells<br>Median (range) | 2.72<br>(0.89-6.63) | 1.51<br>(0.89-2.17) | 2.23<br>(1.37-2.86) | 3.94<br>(1.14-6.63) | 4.29<br>(2.53-5.64) |
| **Plasma cells**<br>%CD45<sup>+</sup> cells<br>Median (range) | 0.20<br>(0.03-0.77) | 0.14<br>(0.12-0.25) | 0.25<br>(0.03-0.77) | 0.16<br>(0.04-0.63) | 0.20<br>(0.10-0.57) |
| **Transitional B cells**<br>%CD45<sup>+</sup> cells<br>Median (range) | 9.48<br>(3.25-24.75) | 14.51<br>(11.22-24.75) | 10.92<br>(3.25-12.71) | 8.06<br>(5.08-13.53) | 6.28<br>(3.52-16.21) |
| **NK cells**<br>%CD45<sup>+</sup> cells<br>Median (range) | 0.63<br>(0.25-1.58) | 0.57<br>(0.42-1.58) | 0.77<br>(0.38-1.26) | 0.63<br>(0.25-1.34) | 0.64<br>(0.35-1.24) |
| **NK-T cells**<br>%CD45<sup>+</sup> cells<br>Median (range) | 0.09<br>(0.02-0.52) | 0.06<br>(0.03-0.07) | 0.08<br>(0.02-0.52) | 0.11<br>(0.05-0.19) | 0.14<br>(0.08-0.23) |
| **CD8 TCM**<br>%CD45<sup>+</sup> cells<br>Median (range) | 0.92<br>(0.22-3.38) | 0.46<br>(0.31-0.60) | 0.92<br>(0.23-3.10) | 1.14<br>(0.22-3.14) | 1.90<br>(0.50-3.38) |
| **CD8 TN**<br>%CD45<sup>+</sup> cells<br>Median (range) | 3.37<br>(0.62-9.69) | 5.19<br>(3.23-8.21) | 2.65<br>(1.75-7.42) | 3.32<br>(1.53-5.90) | 3.75<br>(0.62-9.69) |
| **CD8 TEMRA**<br>%CD45<sup>+</sup> cells<br>Median (range) | 0.36<br>(0.11-2.34) | 0.47<br>(0.2-1.12) | 0.33<br>(0.14-0.79) | 0.48<br>(0.20-1.57) | 0.20<br>(0.11-2.34) |
| **CD8 TEM**<br>%CD45<sup>+</sup> cells<br>Median (range) | 0.83<br>(0.17-3.13) | 0.38<br>(0.17-0.84) | 0.58<br>(0.27-1.62) | 0.92<br>(0.39-2.88) | 1.14<br>(0.65-3.13) |
| **Activated Tregs**<br>%CD45<sup>+</sup> cells<br>Median (range) | 0.56<br>(0.13-1.74) | 0.50<br>(0.27-0.71) | 0.49<br>(0.20-1.13) | 0.62<br>(0.22-1.74) | 0.57<br>(0.13-1.10) |
| **Naive Tregs**<br>%CD45<sup>+</sup> cells<br>Median (range) | 2.62<br>(0.91-7.12) | 2.24<br>(1.31-3.11) | 2.73<br>(1.37-5.33) | 3.21<br>(1.31-7.12) | 2.60<br>(0.91-3.45) |
| **TFH cells**<br>%CD45<sup>+</sup> cells<br>Median (range) | 9.84<br>(0.50-17.32) | 10.95<br>(8.32-17.32) | 10.74<br>(4.59-14.71) | 9.80<br>(4.64-13.24) | 7.34<br>(0.50-14.33) |
| **CD4 Thelper cells**<br>%CD45<sup>+</sup> cells<br>Median (range) | 11.90<br>(1.90-26.26) | 6.51<br>(4.96-9.12) | 8.81<br>(1.90-13.39) | 15.18<br>(7.94-19.69) | 16.78<br>(6.37-26.26) |
| **CD4 TN**<br>%CD45<sup>+</sup> cells<br>Median (range) | 5.58<br>(0.86-16.73) | 6.83<br>(5.06-12.27) | 4.69<br>(3.19-8.66) | 6.41<br>(3.66-9.59) | 5.57<br>(0.86-16.73) |
| **CD4 TEMRA**<br>%CD45<sup>+</sup> cells<br>Median (range) | 0.46<br>(0.07-1.20) | 0.48<br>(0.23-0.75) | 0.51<br>(0.23-1.06) | 0.44<br>(0.21-1.20) | 0.56<br>(0.07-0.94) |
| **Monocytes**<br>%CD45<sup>+</sup> cells<br>Median (range) | 0.36<br>(0.14-1.08) | 0.45<br>(0.21-0.58) | 0.46<br>(0.21-0.86) | 0.34<br>(0.16-1.08) | 0.34<br>(0.14-0.99) |
| **cDCs**<br>%CD45<sup>+</sup> cells<br>Median (range) | 0.23<br>(0.09-0.87) | 0.19<br>(0.17-0.25) | 0.21<br>(0.09-0.87) | 0.26<br>(0.12-0.58) | 0.24<br>(0.14-0.50) |
| **pDCs**<br>%CD45<sup>+</sup> cells<br>Median (range) | 0.83<br>(0.13-2.98) | 0.51<br>(0.42-1.59) | 0.98<br>(0.29-2.07) | 0.68<br>(0.35-1.64) | 1.04<br>(0.13-2.98) |
| **ILCs**<br>%CD45<sup>+</sup> cells<br>Median (range) | 0.34<br>(0.01-1.24) | 0.32<br>(0.26-0.38) | 0.33<br>(0.10-1.24) | 0.44<br>(0.07-0.64) | 0.33<br>(0.01-0.85) |

**Table 6.**
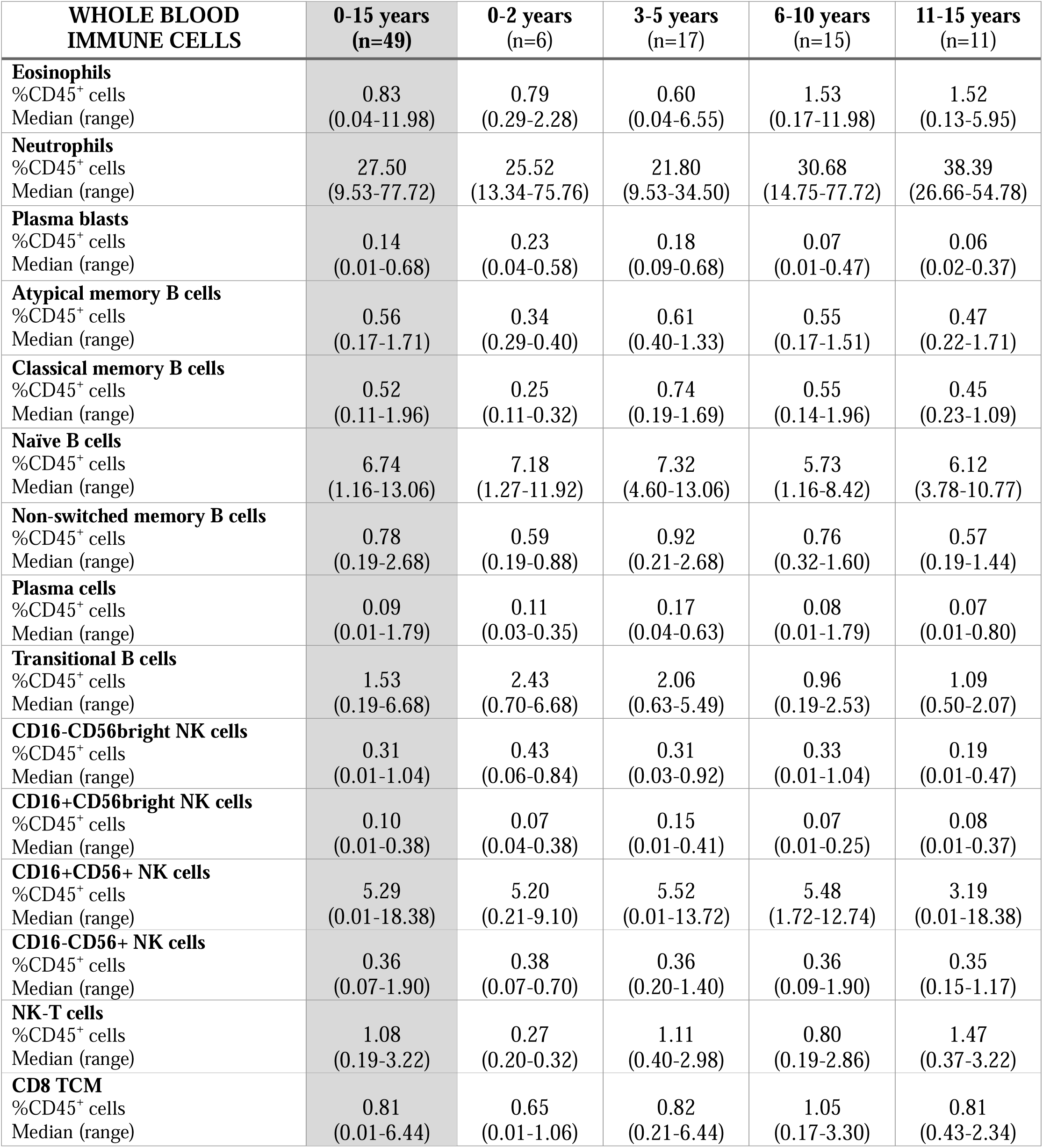
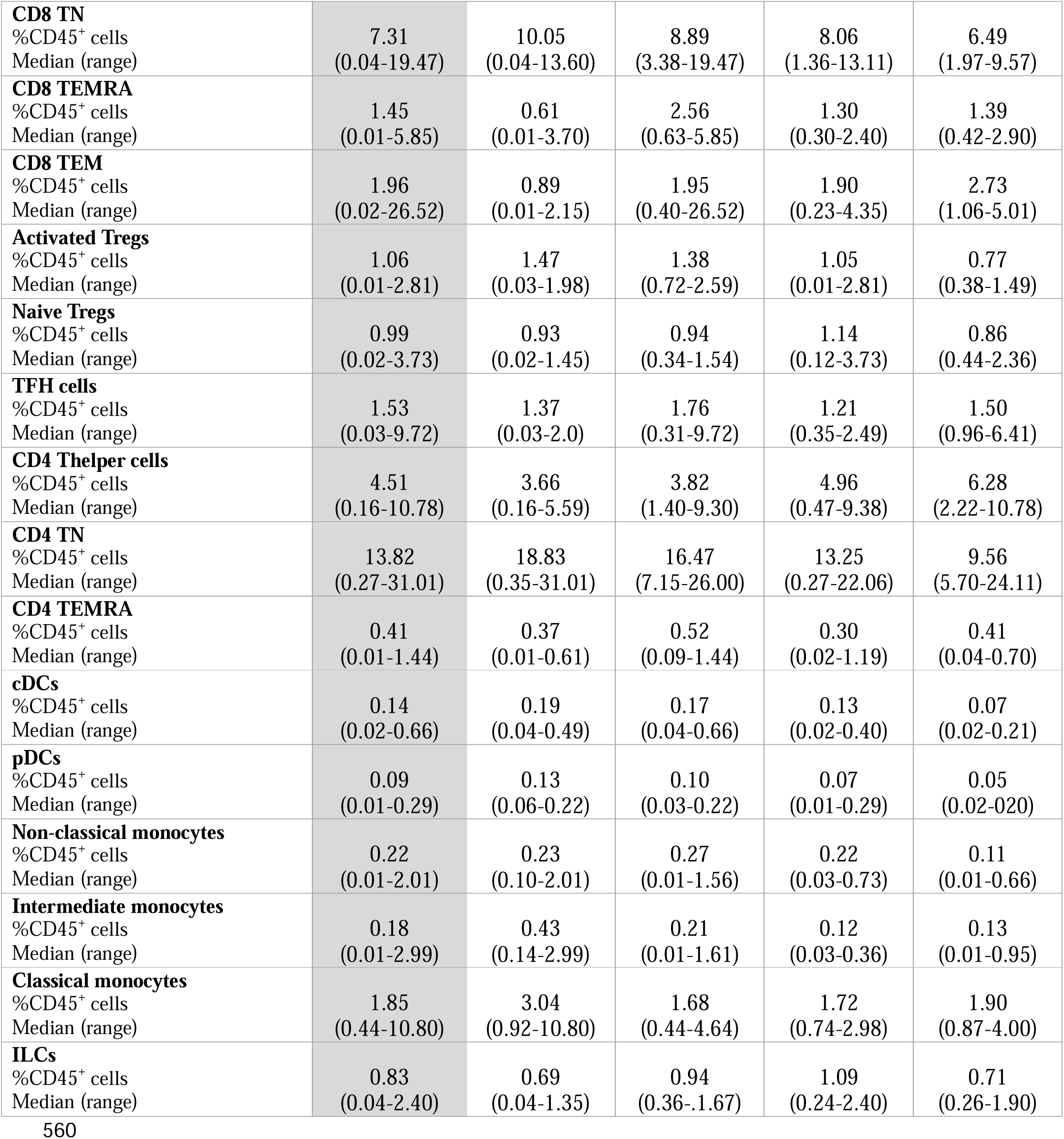
Immune cell subtype frequency in whole blood samples from children aged 0-15 years.

| **WHOLE BLOOD**<br>**IMMUNE CELLS** | **0-15 years**<br>**(n=49)** | **0-2 years**<br>**(n=6)** | **3-5 years**<br>**(n=17)** | **6-10 years**<br>**(n=15)** | **11-15 years**<br>**(n=11)** |
| --- | --- | --- | --- | --- | --- |
| **Eosinophils**<br>%CD45<sup>+</sup> cells<br>Median (range) | 0.83<br>(0.04-11.98) | 0.79<br>(0.29-2.28) | 0.60<br>(0.04-6.55) | 1.53<br>(0.17-11.98) | 1.52<br>(0.13-5.95) |
| **Neutrophils**<br>%CD45<sup>+</sup> cells<br>Median (range) | 27.50<br>(9.53-77.72) | 25.52<br>(13.34-75.76) | 21.80<br>(9.53-34.50) | 30.68<br>(14.75-77.72) | 38.39<br>(26.66-54.78) |
| **Plasma blasts**<br>%CD45<sup>+</sup> cells<br>Median (range) | 0.14<br>(0.01-0.68) | 0.23<br>(0.04-0.58) | 0.18<br>(0.09-0.68) | 0.07<br>(0.01-0.47) | 0.06<br>(0.02-0.37) |
| **Atypical memory B cells**<br>%CD45<sup>+</sup> cells<br>Median (range) | 0.56<br>(0.17-1.71) | 0.34<br>(0.29-0.40) | 0.61<br>(0.40-1.33) | 0.55<br>(0.17-1.51) | 0.47<br>(0.22-1.71) |
| **Classical memory B cells**<br>%CD45<sup>+</sup> cells<br>Median (range) | 0.52<br>(0.11-1.96) | 0.25<br>(0.11-0.32) | 0.74<br>(0.19-1.69) | 0.55<br>(0.14-1.96) | 0.45<br>(0.23-1.09) |
| **Naïve B cells**<br>%CD45<sup>+</sup> cells<br>Median (range) | 6.74<br>(1.16-13.06) | 7.18<br>(1.27-11.92) | 7.32<br>(4.60-13.06) | 5.73<br>(1.16-8.42) | 6.12<br>(3.78-10.77) |
| **Non-switched memory B cells**<br>%CD45<sup>+</sup> cells<br>Median (range) | 0.78<br>(0.19-2.68) | 0.59<br>(0.19-0.88) | 0.92<br>(0.21-2.68) | 0.76<br>(0.32-1.60) | 0.57<br>(0.19-1.44) |
| **Plasma cells**<br>%CD45<sup>+</sup> cells<br>Median (range) | 0.09<br>(0.01-1.79) | 0.11<br>(0.03-0.35) | 0.17<br>(0.04-0.63) | 0.08<br>(0.01-1.79) | 0.07<br>(0.01-0.80) |
| **Transitional B cells**<br>%CD45<sup>+</sup> cells<br>Median (range) | 1.53<br>(0.19-6.68) | 2.43<br>(0.70-6.68) | 2.06<br>(0.63-5.49) | 0.96<br>(0.19-2.53) | 1.09<br>(0.50-2.07) |
| **CD16-CD56bright NK cells**<br>%CD45<sup>+</sup> cells<br>Median (range) | 0.31<br>(0.01-1.04) | 0.43<br>(0.06-0.84) | 0.31<br>(0.03-0.92) | 0.33<br>(0.01-1.04) | 0.19<br>(0.01-0.47) |
| **CD16+CD56bright NK cells**<br>%CD45<sup>+</sup> cells<br>Median (range) | 0.10<br>(0.01-0.38) | 0.07<br>(0.04-0.38) | 0.15<br>(0.01-0.41) | 0.07<br>(0.01-0.25) | 0.08<br>(0.01-0.37) |
| **CD16+CD56+ NK cells**<br>%CD45<sup>+</sup> cells<br>Median (range) | 5.29<br>(0.01-18.38) | 5.20<br>(0.21-9.10) | 5.52<br>(0.01-13.72) | 5.48<br>(1.72-12.74) | 3.19<br>(0.01-18.38) |
| **CD16-CD56+ NK cells**<br>%CD45<sup>+</sup> cells<br>Median (range) | 0.36<br>(0.07-1.90) | 0.38<br>(0.07-0.70) | 0.36<br>(0.20-1.40) | 0.36<br>(0.09-1.90) | 0.35<br>(0.15-1.17) |
| **NK-T cells**<br>%CD45<sup>+</sup> cells<br>Median (range) | 1.08<br>(0.19-3.22) | 0.27<br>(0.20-0.32) | 1.11<br>(0.40-2.98) | 0.80<br>(0.19-2.86) | 1.47<br>(0.37-3.22) |
| **CD8 TCM**<br>%CD45<sup>+</sup> cells<br>Median (range) | 0.81<br>(0.01-6.44) | 0.65<br>(0.01-1.06) | 0.82<br>(0.21-6.44) | 1.05<br>(0.17-3.30) | 0.81<br>(0.43-2.34) |
| **CD8 TN**<br>%CD45<sup>+</sup> cells<br>Median (range) | 7.31<br>(0.04-19.47) | 10.05<br>(0.04-13.60) | 8.89<br>(3.38-19.47) | 8.06<br>(1.36-13.11) | 6.49<br>(1.97-9.57) |
| **CD8 TEMRA**<br>%CD45<sup>+</sup> cells<br>Median (range) | 1.45<br>(0.01-5.85) | 0.61<br>(0.01-3.70) | 2.56<br>(0.63-5.85) | 1.30<br>(0.30-2.40) | 1.39<br>(0.42-2.90) |
| **CD8 TEM**<br>%CD45<sup>+</sup> cells<br>Median (range) | 1.96<br>(0.02-26.52) | 0.89<br>(0.01-2.15) | 1.95<br>(0.40-26.52) | 1.90<br>(0.23-4.35) | 2.73<br>(1.06-5.01) |
| **Activated Tregs**<br>%CD45<sup>+</sup> cells<br>Median (range) | 1.06<br>(0.01-2.81) | 1.47<br>(0.03-1.98) | 1.38<br>(0.72-2.59) | 1.05<br>(0.01-2.81) | 0.77<br>(0.38-1.49) |
| **Naive Tregs**<br>%CD45<sup>+</sup> cells<br>Median (range) | 0.99<br>(0.02-3.73) | 0.93<br>(0.02-1.45) | 0.94<br>(0.34-1.54) | 1.14<br>(0.12-3.73) | 0.86<br>(0.44-2.36) |
| **TFH cells**<br>%CD45<sup>+</sup> cells<br>Median (range) | 1.53<br>(0.03-9.72) | 1.37<br>(0.03-2.0) | 1.76<br>(0.31-9.72) | 1.21<br>(0.35-2.49) | 1.50<br>(0.96-6.41) |
| **CD4 Thelper cells**<br>%CD45<sup>+</sup> cells<br>Median (range) | 4.51<br>(0.16-10.78) | 3.66<br>(0.16-5.59) | 3.82<br>(1.40-9.30) | 4.96<br>(0.47-9.38) | 6.28<br>(2.22-10.78) |
| **CD4 TN**<br>%CD45<sup>+</sup> cells<br>Median (range) | 13.82<br>(0.27-31.01) | 18.83<br>(0.35-31.01) | 16.47<br>(7.15-26.00) | 13.25<br>(0.27-22.06) | 9.56<br>(5.70-24.11) |
| **CD4 TEMRA**<br>%CD45<sup>+</sup> cells<br>Median (range) | 0.41<br>(0.01-1.44) | 0.37<br>(0.01-0.61) | 0.52<br>(0.09-1.44) | 0.30<br>(0.02-1.19) | 0.41<br>(0.04-0.70) |
| **cDCs**<br>%CD45<sup>+</sup> cells<br>Median (range) | 0.14<br>(0.02-0.66) | 0.19<br>(0.04-0.49) | 0.17<br>(0.04-0.66) | 0.13<br>(0.02-0.40) | 0.07<br>(0.02-0.21) |
| **pDCs**<br>%CD45<sup>+</sup> cells<br>Median (range) | 0.09<br>(0.01-0.29) | 0.13<br>(0.06-0.22) | 0.10<br>(0.03-0.22) | 0.07<br>(0.01-0.29) | 0.05<br>(0.02-0.20) |
| **Non-classical monocytes**<br>%CD45<sup>+</sup> cells<br>Median (range) | 0.22<br>(0.01-2.01) | 0.23<br>(0.10-2.01) | 0.27<br>(0.01-1.56) | 0.22<br>(0.03-0.73) | 0.11<br>(0.01-0.66) |
| **Intermediate monocytes**<br>%CD45<sup>+</sup> cells<br>Median (range) | 0.18<br>(0.01-2.99) | 0.43<br>(0.14-2.99) | 0.21<br>(0.01-1.61) | 0.12<br>(0.03-0.36) | 0.13<br>(0.01-0.95) |
| **Classical monocytes**<br>%CD45<sup>+</sup> cells<br>Median (range) | 1.85<br>(0.44-10.80) | 3.04<br>(0.92-10.80) | 1.68<br>(0.44-4.64) | 1.72<br>(0.74-2.98) | 1.90<br>(0.87-4.00) |
| **ILCs**<br>%CD45<sup>+</sup> cells<br>Median (range) | 0.83<br>(0.04-2.40) | 0.69<br>(0.04-1.35) | 0.94<br>(0.36-1.67) | 1.09<br>(0.24-2.40) | 0.71<br>(0.26-1.90) |

### 4. Airway and blood immune cell dynamics over the first 15 years of life

To identify age-specific cellular associations, we performed two-tailed spearman correlation analyses on cell proportion data in each tissue from children aged 1-15 years. No significant age associations for immune cell populations were seen in nasal or alveolar samples after correction for the false discovery rate. In contrast, for bronchial samples, macrophages and NK-T cells increased with age (spearman coefficient (ρ)=0.80, FDR-P=0.002; ρ=0.63, FDR-P=0.03, respectively) while a subset of neutrophils expressing intermediate levels of activation marker CD66b and high levels of CD16 reduced with age (ρ= −0.71, FDR-P=0.01) (Figure 4A). In tonsil samples, transitional B cells decreased with age (ρ= −0.62, FDR-P=0.002), while classical memory B cells, CD8 TEM cells and CD4 Thelper cells increased with age (ρ=0.51, FDR-P=0.02; ρ=0.47, FDR-P=0.04; ρ=0.63, FDR-P=0.002, respectively) (Figure 4B). The same associations with age were also observed in adenoid samples, where transitional B cells decreased with age (ρ= −0.51, FDR-P=0.007) and classical memory B cells, CD8 TEM and CD4 Thelper cells increased with age (ρ=0.43, FDR-P=0.02; ρ=0.52, FDR-P=0.006; ρ=0.62, FDR-P=0.0008, respectively) (Figure 4C). Additionally, four other adenoid immune cells were also associated with age. These were GC B cells and T follicular helper (TFH) cells which decreased with age (ρ= −0.41, FDR-P=0.03; ρ= −0.42, FDR-P=0.02, respectively), and non-switched memory B cells and NK-T cells which increased with age (ρ=0.69, FDR-P=0.0008; ρ=0.62, FDR-P=0.0008, respectively) (Figure 4C). Like the tonsil and adenoid tissue, blood transitional B cells, NK- T cells, and CD4 Thelper cells were also associated with age (ρ= −0.52, FDR-P=0.001; ρ=0.47, FDR-P=0.005; ρ=0.44, FDR-P=0.009, respectively) (Figure 4D). Blood neutrophils increased with age (ρ= 0.40, FDR-P=0.02), while intermediate monocytes and cDCs decreased with age (ρ= −0.42, FDR-P=0.01; ρ= − 0.38, FDR-P=0.02) (Figure 4D). Blood CD4 TN cells decreased with age (ρ=-0.34, FDR-P=0.049) and CD8 TEM cells also tended to increase with age (ρ=0.31, raw-P=0.04, FDR-P=0.09). Individual dot plots depicting correlations for each significant cell type in each tissue are presented in Supplementary Figures 3 and 4.

**Figure 4.**
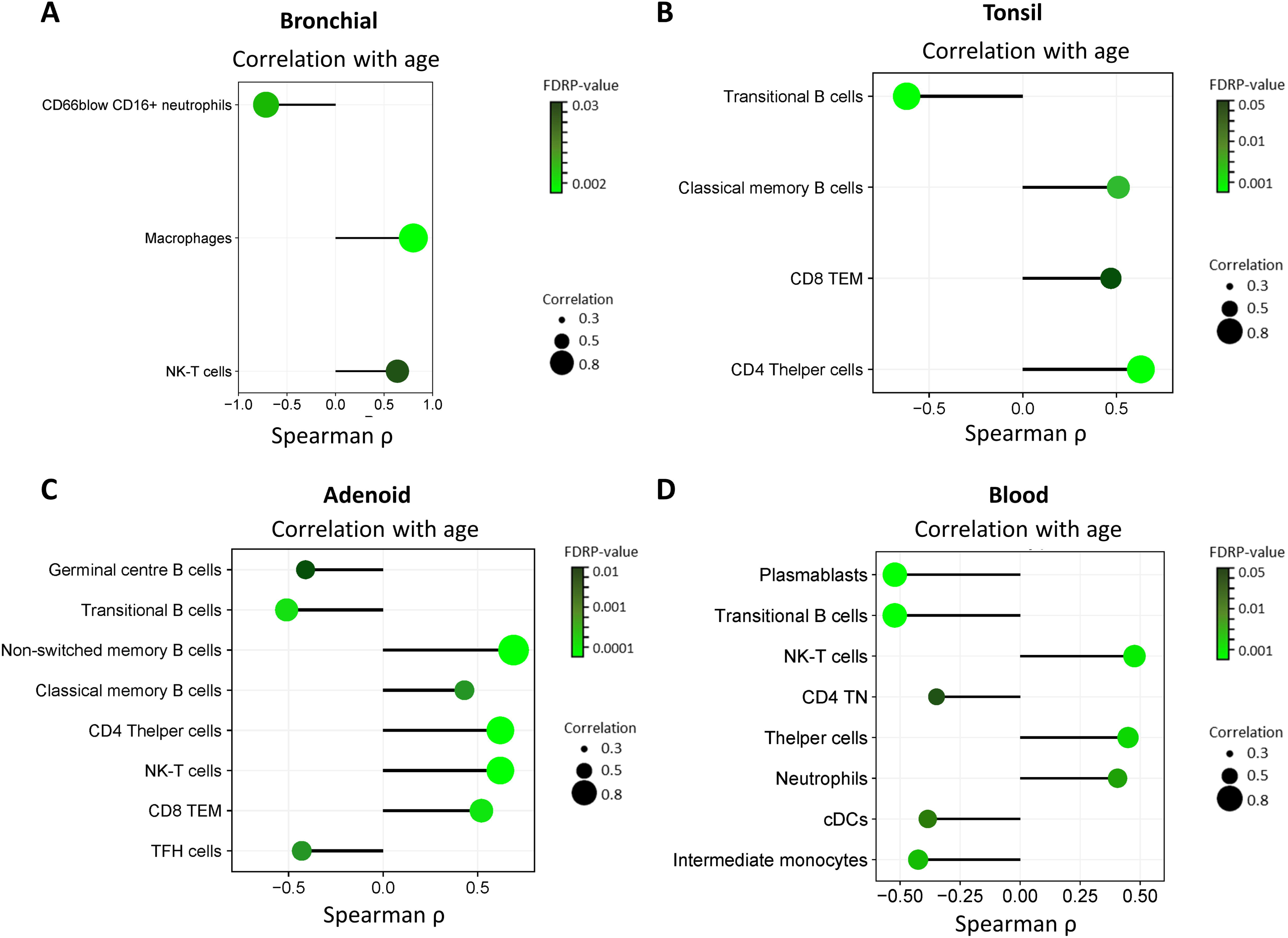
Immune cell type proportions in bronchial, tonsil, adenoid and blood samples are associated with age. **(A)** Spearman coefficient (ρ) and FDR-corrected p-values for nasal immune cell types significantly associated with age: CD66b^low^CD16^+^ neutrophils, macrophages, and NK-T cells. **(B)** Spearman ρ and FDR-corrected p-values for bronchial immune cell types significantly associated with age: transitional B cells, classical memory B cells, CD8 T_EM_, and CD4 Thelper cells. **(C)** Spearman ρ and FDR-corrected p-values for adenoid immune cell types significantly associated with age: germinal centre B cells, transitional B cells, non-switched memory B cells, classical memory B cells, CD4 Thelper cells, NK-T cells, CD8 T_EM_, and TFH cells. **(D)** Spearman ρ and FDR-corrected p-values for blood immune cell types significantly associated with age: plasmablasts, transitional B cells, NK-T cells, CD4 TN cells, CD4 Thelper cells, neutrophils, intermediate monocytes, and cDCs. Note there were no significant correlations with age for cell types in nasal or alveolar samples. Correlation p-values were determined by two-tailed spearman test and corrected for false discovery rate using the Benjamini Hochberg approach. FDR-P<0.05 was considered significant.

These results demonstrate maturation of the airway immune system with age, centred around the development of memory B cells, memory CD8 T cells, and CD4 Teffector populations. Notably, NK-T cell proportions were also highly dynamic, increasing significantly with age in three locations (bronchial, adenoid, and blood) as well as in the alveolar region albeit not significantly when correcting for FDR (ρ=0.88, raw-P=0.007, FDR-P=0.1).

### 5. Cross-tissue analysis of cell type proportions in the upper and lower airway

We next directly compared the composition of immune cells across nasal, bronchi, and alveolar regions. As can be seen in Figure 5A, cell type proportions vary throughout the airway, with granulocytes (neutrophils and eosinophils) more highly abundant in nasal and bronchial tissues; a progressive increase in the proportions of macrophages down the airway; and a shift from CD8 T cell dominance in nasal samples to more equal CD8:CD4 T cell ratios in the alveolar region. Cross-sectional statistical analysis showed that proportions of granulocytes were significantly higher in both the nasal and bronchial samples when compared to alveolar samples (Figure 5B). Macrophage proportions were significantly higher in bronchial compared to nasal, in alveolar compared to nasal, and in alveolar compared to bronchial samples (Figure 5B). Regarding lymphocytes, CD8 T cells and B cells were significantly higher in the nasal than the alveolar samples, and NK-T cells were significantly lower in nasal samples when compared to bronchial samples (Figure 5B). The list of significant cell types and their associated medians and FDR-P values are reported in Supplementary Table 5.

**Figure 5.**
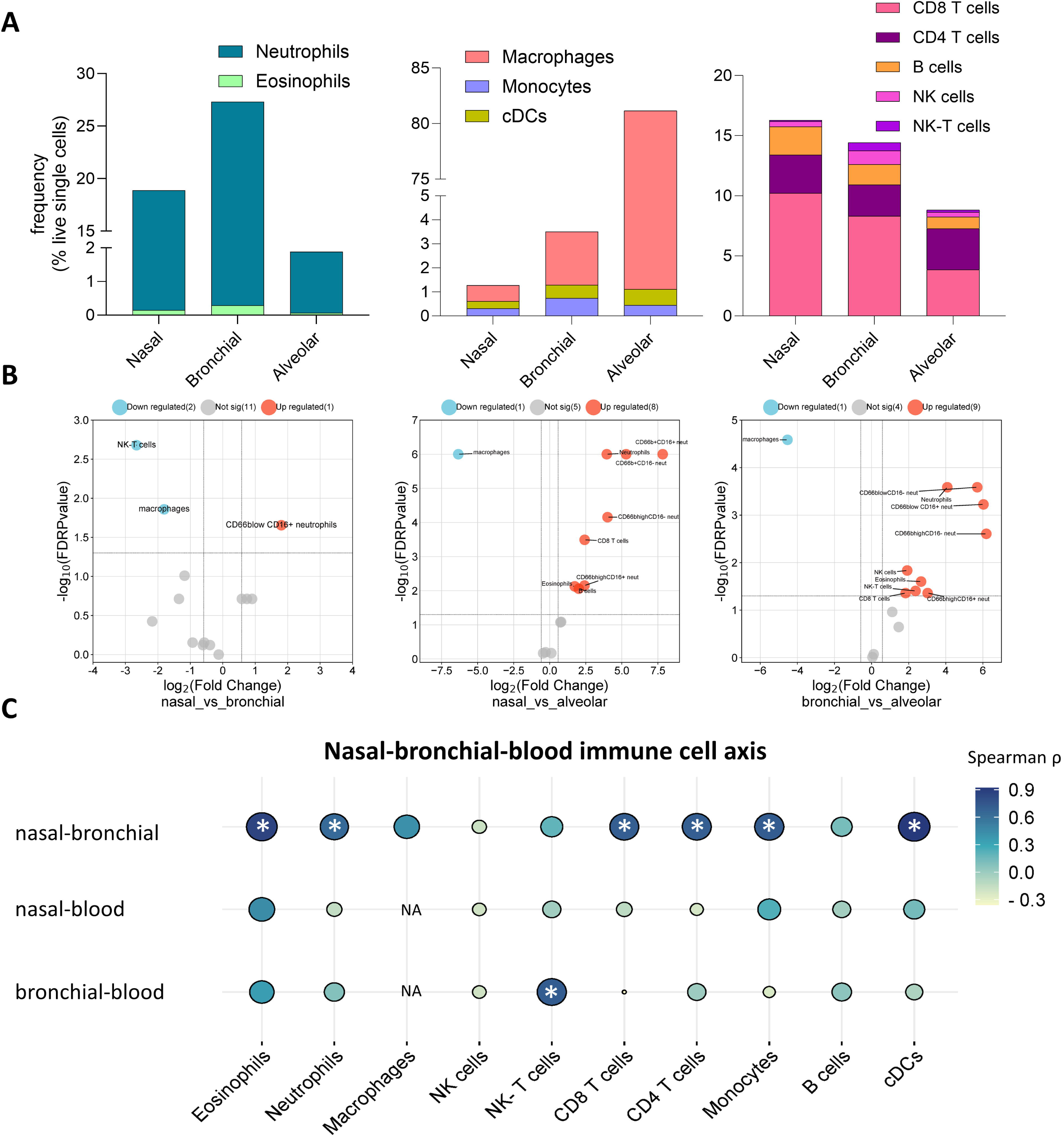
Cross-tissue analysis of cell type proportion in lung and blood compartments reveals high correlation between upper and lower airway brushings but limited correlation between the lung and blood in children. **(A)** Median immune cell type proportion in nasal, bronchial, and alveolar samples for granulocytes (left panel), myeloid cells (centre panel) and lymphocytes (right panel). **(B)** Volcano plots depicting cross-sectional statistical analysis of cell types in nasal_vs_bronchial samples (left panel), nasal_vs_alveolar samples (centre panel), and bronchial_vs_alveolar samples (right panel). **(C)** Ballon plot depicting intra- individual correlations of cell type proportions for participants who had nasal, bronchial, and blood samples analysed. * Depicts significant correlation. NA: Not Applicable, as macrophages are not present in blood. Cross-sectional p-values (part B) were determined by Mann-Whitney U-test and corrected for false discovery rate using the Benjamini Hochberg approach. FDR p<0.05 and fold change >1.5 was considered significant. Intra-individual correlation p-values (part C) were determined by two-tailed spearman test and corrected for false discovery rate using the Benjamini Hochberg approach. FDR-P<0.05 was considered significant.

As we had collected multiple sample types per participant, we were able to test for correlations in immune cell proportions between the upper and lower airway in the same individuals. Due to limited numbers of paired BAL-nasal, and paired BAL-bronchial samples, we could not perform this analysis for these sample types. However, we were able to perform cross-tissue analysis for paired nasal and bronchial samples (n=13), paired nasal and blood samples (n=29) and paired bronchial and blood samples (n=15). This revealed that nasal and bronchial samples were highly correlated within individuals, with positive correlations observed between eosinophils (spearman coefficient (ρ)=0.85, FDR-P=0.002), neutrophils (ρ=0.64, FDR-P=0.03), CD4 T cells (ρ=0.70, FDR-P=0.02), CD8 T cells (ρ=0.69, FDR-P=0.02), monocytes (ρ=0.72, FDR-P=0.02) and cDCs (ρ=0.92, FDR-P=0.0002) in the nasal and bronchial samples (Figure 5C). On the contrary, we observed no statistical correlation in immune cell proportions between blood and nasal samples within individuals, and only NK-T cells were significantly correlated between blood and bronchial samples (ρ=0.72, FDR-P=0.03) (Figure 5C). All correlation coefficients and FDR-P values are reported in Supplementary Table 6.

### 6. Cross-tissue analysis of cell type proportions in the upper airway lymphoid tissue

Next, we directly compared the composition of immune cells across tonsil, adenoid, and blood compartments. As can be seen in Figure 6A, tonsil and adenoid cell compositions were similar, particularly for NK-T cells, innate lymphoid cells (ILCs), all CD8 T cell subsets, all CD4 T cell subsets and Tregulatory cells. Blood samples showed higher frequencies of innate immune cells, NK/ILCs, as well as naïve CD8- and CD4- T cells, and lower frequencies of TFH cells and the B cell lineage when compared to tonsils and adenoids (Figure 6A, statistical comparison presented in Supplementary Figure 5). Cross-sectional statistical comparison of tonsils vs adenoids showed that plasmablasts, GC B cells, and neutrophils were significantly higher in tonsils compared to adenoids, while pDCs, NK cells, classical memory B cells, and non-switched memory B cells were significantly higher in adenoids compared to tonsils (Figure 6B). The list of significant cell types and their associated medians and FDR-P values are reported in Supplementary Table 7.

**Figure 6.**
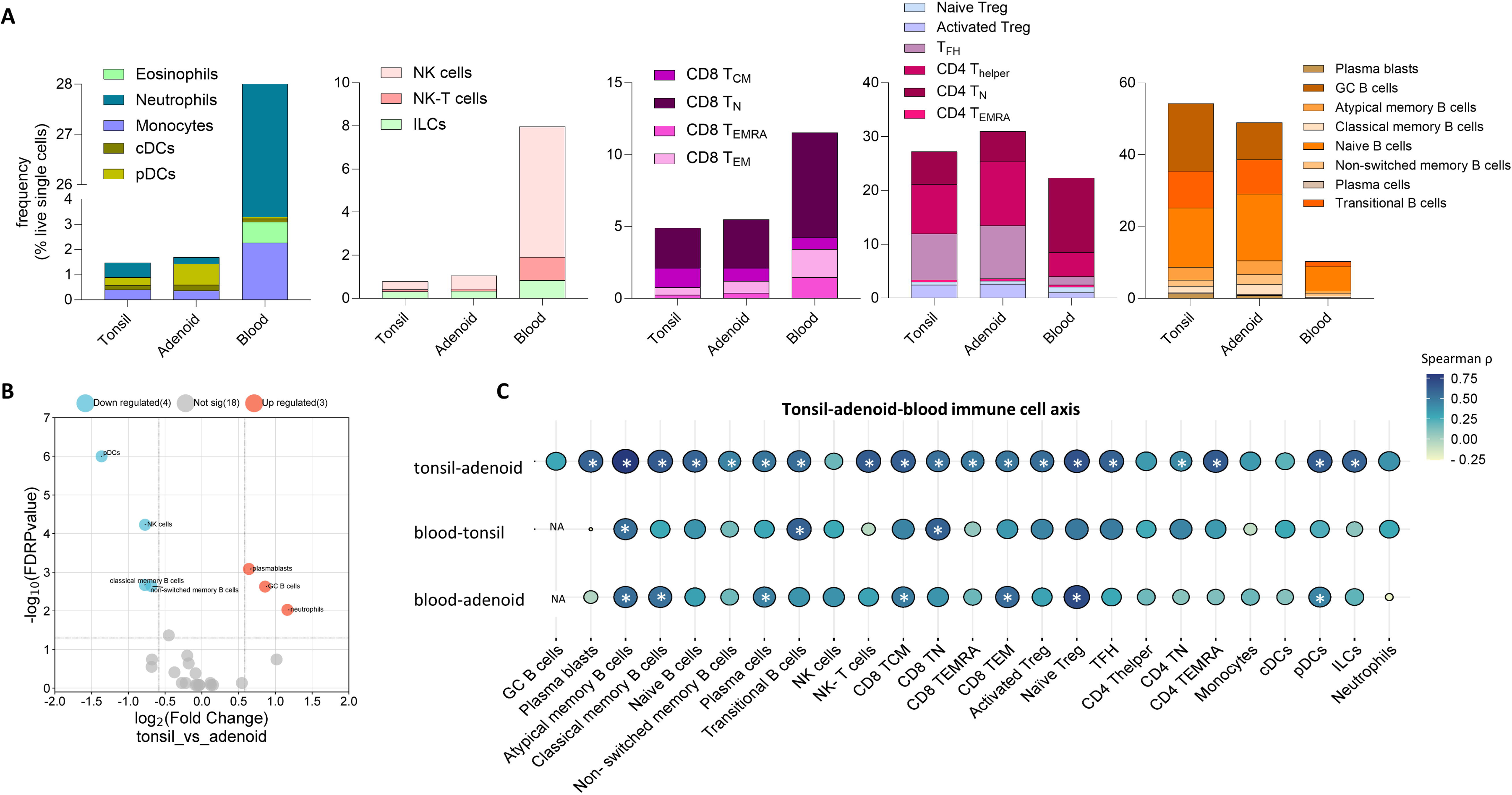
Cross-tissue analysis of cell type proportion in tonsil, adenoid and blood compartments reveals sample-type specific immune cell profiles and an intra-individual tonsil-adenoid-blood immune cell axis in children. **(A)** Median immune cell type proportion in tonsil, adenoid, and blood samples for granulocytes and myeloid cells (first panel), innate lymphocytes (second panel), CD8 T cell populations (third panel), CD4 T cell populations (fourth panel) and B cell populations (fifth panel). **(B)** Volcano plot depicting cross-sectional statistical analysis of cell types in tonsil_vs_adenoid samples. **(C)** Ballon plot depicting intra- individual correlations of cell type proportions for participants who had tonsil, adenoid, and blood samples analysed. * Depicts significant correlation. NA: Not Applicable, as GC B cells are not present in blood. Cross-sectional p-values (part B) were determined by Mann- Whitney U-test and corrected for false discovery rate using the Benjamini Hochberg approach. FDR p<0.05 and fold change >1.5 was considered significant. Intra-individual correlation p-values (part C) were determined by two-tailed spearman test and corrected for false discovery rate using the Benjamini Hochberg approach. FDR-P<0.05 was considered significant.

Finally, we tested for correlations in immune cell proportions between tonsil, adenoid, and blood compartments in paired samples collected from the same individuals (Figure 6C). There were 27 children with paired tonsil and adenoid samples, 25 children with paired tonsil and blood samples, and 30 children with paired adenoid and blood samples. This cross-tissue analysis revealed that tonsil and adenoid samples were highly correlated within individuals, with positive associations observed between 19/25 immune cell populations identified in the lymphoid tissues (Figure 6C). Similarly, blood cells also correlated with cells in the upper airway lymphoid tissue, especially evident for the adenoids where 7/24 populations were positively correlated (Figure 6C). Blood cell proportions positively correlated with tonsil cell proportions for atypical memory B cells, transitional B cells, and CD8 TN within individuals. Blood cell proportions positively correlated with adenoid cell proportions for atypical memory B cells, classical memory B cells, plasma cells, CD8 TCM, CD8 TEM, naïve Tregs and pDCs within individuals. All correlation coefficients and associated FDR-P values are reported in Supplementary Table 8.

## DISCUSSION

In recent years, there has been an explosion in the development of single-cell sequencing based tissue atlases, including a global consortium dedicated to profiling the lung ^8^. These studies have provided an unprecedented level of resolution into gene expression of individual cells across many populations. However, due to the extensive laboratory and specialist computational pipelines required for analysis of single-cell sequencing data, these tools are many years away from application in clinical laboratories. Flow cytometry is routinely used in the diagnosis and assessment of human disease globally, and permits multi-dimensional, high-throughput quantification of millions of cells without the requirement for specialist computational analysis ^9^. To our knowledge, we have developed the first flow cytometry-based reference for immune cells in the airways of children. This includes detailed assessment of cell populations in the nose, upper airway lymphoid tissue, as well as the lower airway epithelium and alveolar space; a detailed analysis of surface markers present on each cell population in each tissue; and investigation of cell frequencies across time and space. We also present a reference for immune cells in whole blood samples collected from the same children. The goal of this work was to understand the relationship between airway and systemic immunity from paired paediatric samples, however we also hope it adds valuable data to other efforts investigating immune development in blood, including fresh whole blood ^10^, cryopreserved whole blood ^11^, and cryopreserved PBMC ^12^ samples. To facilitate the application and implementation of our tissue references, all protocols, flow cytometry panels, and data are openly available.

Our results show tissue-specific maturation of the airway immune system with age. For example, we did not show changes in immune cell proportions throughout childhood in nasal samples, however a marked increase in macrophages and NK-T cells were observed during childhood in bronchial samples. We were fascinated to find strikingly high immune:epithelial cell ratios in nasal and bronchial brushings samples, particularly the high proportions of neutrophils. A recent study that used single-cell sequencing to profile immune cells in nasal swabs from children with SARS-CoV-2 infection as well as non-infected controls reported similar findings. They showed that while immune cells were rarely detected in nasal swabs from healthy adults, samples from healthy children contained high amounts of immune cell subsets with an overall dominance of neutrophils ^13^. Regarding the lymphoid tissues, both tonsil and adenoid samples showed increases in classical memory B cells, CD8 TEM cells and CD4 Thelper cells with age, as well as a decrease in transitional B cells. Adenoid cells, however, appeared more influenced by age, with additional significant associations observed in germinal centre B cells, non-switched memory B cells, NK-T cells and TFH cells. Similar age-related changes in airway lymphoid tissues have been observed previously, particularly related to germinal centre development in tonsils ^14^. For blood immune cells, we replicate what has been observed in another study focusing on lymphocyte populations from a large number of paediatric whole blood samples where plasmablasts, transitional B cells, and CD4 T naïve cells decreased with age and CD4 Thelper cells increased with age ^10^. We additionally show that whole blood NK-T cells, neutrophils, cDCs, and intermediate monocytes are also associated with age. Collectively, these age-related changes in immune cell composition further highlight the importance of developing age-specific references for the airways and blood of children. This work also highlights that many single-cell sequencing based tissue atlases that primarily consist of samples obtained from adults may not be applicable to children, and we support the efforts of the Pediatric Cell Atlas to include data that samples the complete human lifespan ^15^.

Our intra-individual cross-tissue analysis revealed high correlation in immune cell proportions between tissues, particularly for paired nasal and bronchial samples, paired tonsil and adenoid samples, and paired adenoid and blood samples. These findings suggest that easily accessible nasal samples could be used as a surrogate for assessment of immunity in the lower airway of children. If validated in future studies, this could have important implications for clinical testing, and potentially prevent risks associated with general anaesthesia to sample the lower airway. The remarkable correlation in immune cell composition between the tonsils and adenoids is likely explained by the fact both tissues are part of Waldeyer’s ring of pharyngeal lymphoid organs.

We acknowledge the limitations of our dataset, in particular the limited number of BAL samples which did not permit age-related or cross-tissue analysis for this sample type. This is due to the difficulties in obtaining these lower airway samples from children, as well as limitations in sample volumes for research testing. However, we were able to obtain samples from a larger number of children for all other sample types (n=17-49), and these numbers are similar to or above that of other immunological studies using paediatric respiratory specimens ^13, 16–18^. We also acknowledge that these samples were collected as part of clinically indicated procedures, which means that not all samples can be considered ‘healthy’. The main indication in our cohort was OSA, with >80% of samples obtained from children exhibiting or undergoing investigation for this indication. While this may contribute to some variation in tonsil or adenoid immune cell proportions ^18^, the effect of this underlying condition on results obtained from distal sample types in these children (nasal brushings, bronchial brushings, blood) is unclear. A recent review revealed conflicting evidence of immune cell alterations in adult and paediatric OSA, with some studies reporting an inflammatory effect while others reported normal immune responses in these patients ^19^. All children in our cohort were clinically well at the time of sample collection and there was limited evidence of wheezing or respiratory infection in the week leading up to sample collection. Parent questionnaires revealed that the majority of children in our cohort had never been prescribed respiratory medications, and of those prescribed, rates of medication use were similar to what is observed in the Australian community for children aged 0-15years^20^. Likewise, rates of conditions including general atopy, asthma, and eczema, were similar to a recent population-based study of Australian children ^21^. As general anaesthesia is required to collect many of the sample types included in our study, creating control respiratory datasets from healthy children in the community is not feasible, especially datasets that include multiple sample types from the same individual, Our reference can be used as control data for clinical samples collected from children with acute (e.g. infectious), rare (e.g. cystic fibrosis, primary ciliary dyskinesia) and chronic (e.g. suppurative lung disease) respiratory conditions.

In summary, this study advances our knowledge of airway immunity from infancy through to adolescence and provides an easily adaptable, publicly available control dataset to aid in interpretation of clinical samples obtained from children with respiratory disease.

## MATERIALS AND METHODS

### Study participants

This study took place at the Royal Children’s Hospital (RCH, Melbourne Australia) and involved analysis of 178 samples from 66 children (Figure 1A, Supplementary Table 1) aged between 1 and 15 years. Samples were collected at the time of clinically indicated procedures (bronchoscopy, tonsillectomy and/or adenoidectomy). Reasons for procedures are detailed in Supplementary Table 1 and medical history from parent questionnaires are summarised in Supplementary Table 2.

### Collection and processing of samples

Samples were collected at the time of clinically indicated procedures (bronchoscopy, tonsillectomy and/or adenoidectomy). For palatine tonsils (“tonsils”) and adenotonsils (“adenoids’), tissues were removed by the operating team as per standard operating procedures, suspended in RPMI media supplemented with 2% fetal calf serum (FCS), and transferred on ice to the laboratory immediately upon collection. For nasal brushings, samples were collected by a respiratory physician using a 3mm cytology brush at the time of the procedure, suspended in cold RPMI media supplemented with 2% FCS and transferred on ice to the laboratory immediately upon collection. Bronchial brushings were collected by a respiratory physician using a 2mm cytology brush and suspended in cold RPMI media supplemented with 2% FCS and transferred on ice to the laboratory immediately upon collection. Bronchoscopy and bronchoalveolar lavage (BAL) were performed as previously described where indicated for investigation of stridor ^22^. This study used BAL from aliquots two and three from the right middle lobe. BAL samples were transferred on ice to the laboratory immediately upon collection. Venous blood was collected in EDTA tubes at the time of the procedure and transferred to the laboratory at room temperature immediately upon collection. Multiple sample types were collected from each individual (Figure 1A).

Detailed protocols for the collection, processing and flow cytometry analysis of each sample type are openly available at: protocols.io/workspaces/earlyAIR. Briefly, cells were dislodged off nasal and bronchial brushes into media by pipetting for at least 1 minute. Cells were filtered through a 70µM cell strainer into a new 15mL tube and centrifuged at 300xg for 7 minutes at 4 degrees. Cells were counted using the LUNA-FL Dual Fluorescence cell counter and washed once more before 300,000 cells were aliquoted for flow cytometry as described below. BAL samples were centrifuged at 300xg for 7 minutes at 4 degrees. The cell-free supernatant was removed and cells were resuspended in 10mL of media. Cells were filtered through a 70µM cell strainer into a new 15mL tube and centrifuged at 300xg for 7 minutes at 4 degrees. Cells were counted using the LUNA-FL cell counter and washed once more before 300,000 cells were aliquoted for flow cytometry as described below. Tonsil and adenoid samples were processed in glass culture plates with 10mL of media. First, visible blood clots, fat and connective tissue were removed using forceps and scissors. Next, the tissues were minced into a fine paste using scissors in a fresh glass plate with fresh media and filtered through a 100µm cell strainer into a 50mL tube. Cells were centrifuged at 400xg for 5 minutes at room temperature and resuspended in 8mL of fresh media prior to layering onto Ficoll plaque plus as per the manufacturer’s instructions. Cells were centrifuged at 400xg for 30 minutes at room temperature with a slow acceleration and no brake. Cells at the interface between media and the Ficoll solution were collected into a new 15mL tube and washed with media before counting using the LUNA-FL cell counter. Due to the large number of cells collected, cells were diluted between 1:10-1:100 prior to counting. Finally, 500,000 cells were aliquoted for flow cytometry as described below.

### Flow cytometry of nasal brushings, bronchial brushings, and BAL samples

Nasal, bronchial and BAL cells were resuspended in PBS for viability staining using near infra-red viability dye according to manufacturers’ instructions. The viability dye reaction was stopped by the addition of FACS buffer (2% heat-inactivated FCS in PBS) and cells were centrifuged at 400xg for 5 mins 4°C. Cells were resuspended in 25µL human FC-block (diluted 1:10) for 5 minutes at room temperature. The flow cytometry staining panel described in Supplementary Table 3 (made up at 2X concentration) was added 1:1 with the cells and incubated for 30 minutes on ice. Following staining, cells were washed with 1mL FACS buffer and centrifuged at 400xg for 5 minutes 4°C. Cells were resuspended in 150µL FACS buffer for acquisition using the Cytek 5L Aurora. The gating strategy used to define cell types obtained with this panel is depicted in Supplementary Figure 1.

### Flow cytometry of tonsils, adenoids, and whole blood samples

Tonsil, adenoid and lysed whole blood cells were resuspended in PBS for viability staining using near UV-blue viability dye according to manufacturers’ instructions. The viability dye reaction was stopped by the addition of FACS buffer and cells were centrifuged at 400xg for 5 mins 4°C. Cells were resuspended in 40µL human FC-block (diluted 1:10) for 5 minutes at room temperature, following which 10µL of Brilliant Stain Buffer Plus was added. The chemokine panel described in Supplementary Table 4A (made up at 3X concentration) was added at 25µL per sample and cells incubated for 10 minutes at room temperature in the dark. The panel described in Supplementary Table 4B (made up at 3X concentration) was added at 50µL per sample and cells incubated for 30 minutes at room temperature in the dark. Following staining, cells were washed with 1mL FACS buffer and centrifuged at 400xg for 5 minutes 4°C. Cells were resuspended in 150µL FACS buffer for acquisition using the Cytek 5L Aurora. The gating strategy used to define cell types obtained with this panel is depicted in Supplementary Figure 2. The panel described in Supplementary Table 4 is a modified version of the published OMIP-069 ^23^.

### Data analysis

Gating of flow cytometry data was performed in FlowJo version 10.9.0. As samples were acquired fresh on the day of collection over multiple batches, all data was normalised using the cyCombine plugin ^24^. For each tissue type, cell populations were annotated based on expression of known lineage markers included in each panel (see gating strategies for each tissue in the Supplement). Cell population frequencies were expressed as a proportion of live, single cells or as a proportion of live, single, CD45^+^ cells when referring to analysis of immune cell types only. For age-specific immune cell reference ranges (Tables 1-6), median frequencies (% of live, single, CD45^+^ cells) were calculated for each cell type within each age group (0-2years, 3-5 years, 6-10 years, 11-15 years) and the ranges also reported. For unsupervised analyses, a single concatenated file was created for each tissue type, containing 10,000 downsampled cells from each participant. These concatenated files were used for Uniform Manifold Approximation and Projection (UMAP) analysis (using the default settings of the UMAP plugin for FlowJo) and to calculate the median fluorescence intensity (MFI) of each lineage marker for each cell type in each tissue. Hierarchical clustering analysis and heatmap visualisation of the MFI data was done using the pheatmap package in R Studio version 4.3.0. For cross-sectional statistical analysis, cell proportions were compared using Mann Whitney U-tests. All p-values were corrected for false discovery rate using the Benjamini and Hochberg approach ^25^. An FDR corrected p<0.05 and a fold change of >1.5 was required to reach statistical significance. Age and intra-individual correlations were determined using two-tailed spearman tests. Spearman P-values were corrected for false discovery rate using the Benjamini and Hochberg approach. An FDR corrected p<0.05 was required to reach statistical significance.

## Ethics statement

The studies involving human participants were reviewed and approved by Royal Children’s Hospital Human Research Ethics Committee (HREC #25054 and # 88144). Written informed consent to participate in the studies was provided by the participants’ legal guardian/next of kin.

## Supporting information

Data repository

## Data Availability

All data produced in the present work are contained in the manuscript

https://www.protocols.io/workspaces/earlyAIR

## ACKNOWLEDGEMENTS

This work was funded by a Chan Zuckerberg Initiative Single-Cell Biology Grant (2021- 237883). We thank the children and parents who participated in our studies, without whom this work would not exist.

