## Supplemental file for "A cross-tissue, age-specific flow cytometry reference for immune cells in the airways and blood of children"

**Supplementary Table 1.** Demographics and characteristics of the study cohort.

| | **Nasal brushings** | **Bronchial brushings** | **BAL** | **Adenoids** | **Tonsils** | **Whole blood** |
|---|---|---|---|---|---|---|
| **Total number** | 37 | 17 | 8 | 36 | 31 | 49 |
| **Age: years, median (range)** | 4.69 (1.1-13.2) | 6.18 (2.3-12.1) | 3.03 (0.5-10.3) | 5.5 (1.6-15.2) | 4.7 (1.1-13.2) | 6.65 (1.1-15.2) |
| **Sex: male, n (%)** | 21 (56.7) | 11(64.7) | 5 (62.5) | 21 (58.3) | 18 (58.1) | 29 (59) |
| **Reason for procedure [n, (%)]** | | | | | | |
| - OSA | 32 (86.5) | 16 (94.1) | - | 33 (91.7) | 30 (96.8) | 40 (81.6) |
| -hearing concerns | 1 (2.7) | - | - | - | - | 3 (6.1) |
| -history of tonsillitis | 1 (2.7) | - | - | 1 (2.8) | 1 (3.2) | 1 (2.0) |
| -investigation of stridor | 3 (8.1) | - | 8 (100) | - | - | 3 (6.1) |
| -nasal obstruction | - | 1 (5.9) | - | 2 (5.5) | - | 1 (2.0) |
| -vocal cord polyp | - | - | - | - | - | 1 (2.0) |

OSA – obstructive sleep apnoea, BAL – bronchoalveolar lavage.

**Supplementary Table 2.** Summary data from parent questionnaires for medical history, respiratory status and respiratory medications for the whole cohort (n=66 children).

| **Cohort summary statistics** | No (%) | Yes (%) | parent unsure or data missing (%) | Other information |
|---|---|---|---|---|
| **Medical history** | | | | |
| Bronchiolitis_hospital_ever | 75.8 | 13.6 | 10.6 | 78% of "yes" was >1 year ago |
| Asthma_hospital_ever | 78.8 | 9.1 | 12.1 | 50% of "yes" was >1 year ago |
| Croup_hospital_ever | 83.3 | 7.6 | 9.1 | 80% of "yes" was >1 year ago |
| Lower respiratory tract infection_hospital_ever | 83.3 | 7.6 | 9.1 | 80% of "yes" was >1 year ago |
| Upper respiratory tract infection_hospital_ever | 83.3 | 6.1 | 10.6 | 50% of "yes" was >1 year ago |
| Pneumonia_hospital_ever | 86.4 | 3 | 10.6 | 50% of "yes" was >1 year ago |
| Rhinovirus_hospital_ever | 86.4 | 1.5 | 12.1 | 100% of "yes" was >1 year ago |
| Influenza_hospital_ever | 89.4 | 0 | 10.6 | NA |
| Doctor_diagnosed_asthma_ever | 65.1 | 25.8 | 9.1 | NA |
| Doctor_diagnosed_allergic_rhinitis | 54.5 | 36.4 | 9.1 | NA |
| Doctor_diagnosed_protracted_bacterial_bronchitis_ever | 84.8 | 6.1 | 9.1 | NA |
| Doctor_confirmed_eczema_ever | 62.1 | 27.3 | 10.6 | NA |
| Doctor_confirmed_allergies (food, dust, plant, animal) | 66.7 | 21.2 | 12.1 | NA |
| **Current respiratory status** | | | | |
| Cough_last_7days | 77.3 | 13.6 | 9.1 | NA |
| Cough_last_month | 57.6 | 33.3 | 9.1 | NA |
| Wheeze_last_7days | 83.3 | 7.6 | 9.1 | NA |
| Wheeze_lastmonth | 74.2 | 16.7 | 9.1 | NA |
| Respiratory_tract_infection_last_7days | 89.4 | 1.5 | 9.1 | NA |
| Respiratory_tract_infection_past_month | 71.2 | 19.7 | 9.1 | NA |
| **Respiratory medications** | | | | |
| Bronchodilator_use_ever | 63.6 | 27.3 | 9.1 | 16.6% of "yes" was >1 year ago |
| Inhaled_steroid_use_ever | 84.8 | 6.1 | 9.1 | 25% of "yes" was >1 year ago |
| Oral_steroid_use_ever | 74.2 | 16.7 | 9.1 | 100% of "yes" was >1 year ago |
| Nasal_steroid_use_ever | 60.9 | 30 | 9.1 | 60% of "yes" was >1 year ago |
| Combination_treatment_use_ever | 86.4 | 4.5 | 9.1 | 100% of "yes" was "as needed" |
| Leukotreine_antagonist_use_ever | 86.4 | 4.5 | 9.1 | 100% of "yes" was "daily" |

**Supplementary Table 3.** Flow cytometry panel for assessment of immune cell populations in nasal brushings, bronchial brushings, and bronchoalveolar lavage (BAL).

| Antibody | Fluorophore | Volume in µL (2X in 100µL) | Supplier | Clone |
|---|---|---|---|---|
| CD16 | BUV395 | 0.5 | Becton Dickinson | 3G8 |
| SIGLEC8 | BUV661 | 1 | Becton Dickinson | 837535 |
| CD56 | BUV737 | 2 | Becton Dickinson | NCAM16.2 |
| CD11b | BUV805 | 2 | Becton Dickinson | D12 |
| CD206 | BV421 | 1 | Becton Dickinson | 19.2 |
| HLADR | V500 | 1 | Becton Dickinson | G46-6 |
| CD19 | BV605 | 2 | Becton Dickinson | SJ25C1 |
| CD8 | BV650 | 1 | Becton Dickinson | RPA-T8 |
| CD45 | BV711 | 2 | Becton Dickinson | HI30 |
| CD14 | BV786 | 4 | Becton Dickinson | M5E2 |
| EPCAM | BB515 | 1 | Becton Dickinson | EBA-1 |
| CD3 | FITC | 2 | Becton Dickinson | SK7 |
| CD66b | PE | 0.5 | Becton Dickinson | G10F5 |
| CD15 | PE-CF594 | 1 | Becton Dickinson | W6D3 |
| CD11c | PE-Cy7 | 2 | Becton Dickinson | B-ly6 |
| CD4 | A700 | 2 | Becton Dickinson | RPA-T4 |
| Live/Dead | NIR | N/A | Invitrogen (Thermo Fisher Scientific) | N/A |

**Supplementary Table 4A.** Flow cytometry panel for assessment of immune cell populations in tonsils, adenoids, and whole blood – chemokines and receptors.

| Antibody | Fluorophore | Volume in µL (3X in 100µL) | Supplier | Clone |
|---|---|---|---|---|
| CCR4 | BUV661 | 3 | Becton Dickinson | 1G1 |
| CCR7 | BV421 | 6 | BioLegend | G043H7 |
| CXCR5 | BV750 | 6 | Becton Dickinson | RF8B2 |
| CXCR3 | PE-Cy7 | 0.75 | BioLegend | G025H7 |

**Supplementary Table 4B.** Flow cytometry panel for assessment of immune cell populations in tonsils, adenoids, and whole blood – all other antibodies.

| Antibody | Fluorophore | Volume in µL (3X in 100µL) | Supplier | Clone |
|---|---|---|---|---|
| CD45RA | BUV395 | 1.5 | Becton Dickinson | 5H9 |
| CD16 | BUV496 | 0.75 | Becton Dickinson | 3G8 |
| CD206 | BUV563 | 1.5 | Becton Dickinson | 19.2 |
| CD56 | BUV737 | 1.5 | Becton Dickinson | NCAM16.2 |
| CD8 | BUV805 | 0.75 | Becton Dickinson | SK1 |
| CD123 | superbright 436 | 3 | Invitrogen (Thermo Fisher Scientific) | 6H6 |
| CD11c | eFluor 450 | 1.5 | Invitrogen (Thermo Fisher Scientific) | 3.9 |
| IgD | BV480 | 0.75 | Becton Dickinson | IA6-2 |
| CD3 | BV510 | 1.5 | BioLegend | SK7 |
| CD20 | Pacific orange | 1.5 | Invitrogen (Thermo Fisher Scientific) | HI47 |
| IgG | BV605 | 6 | Becton Dickinson | G18-145 |
| HLADR | BV650 | 0.75 | Becton Dickinson | G46-6 |
| PD-1 | BV785 | 1.5 | BioLegend | EH12.2H7 |
| CD141 | BB515 | 1.5 | Becton Dickinson | 1A4 |
| CD69 | FITC | 3 | BioLegend | FN50 |
| CD14 | Spark Blue 550 | 1.5 | BioLegend | 63D3 |
| CD45 | PerCP | 3 | BioLegend | 14C2A07 |
| CD4 | cFluor YG584 | 1.5 | Cytek Biosciences | SK3 |
| CD15 | PE-CF594 | 1.5 | Becton Dickinson | W6D3 |
| CD24 | PE-Alexa Fluor 610 | 3 | Invitrogen (Thermo Fisher Scientific) | SN3 |
| CD25 | PE-Alexa Fluor 700 | 3 | Invitrogen (Thermo Fisher Scientific) | CD25-3G10 |
| CD1c | AF647 | 12 | BioLegend | L161 |
| CD19 | Spark NIR 685 | 3 | BioLegend | HIB19 |
| CD127 | APC-R700 | 3 | Becton Dickinson | HIL-7R-M21 |
| CD27 | APC-H7 | 6 | Becton Dickinson | M-T271 |
| CD38 | APC-Fire 810 | 0.75 | BioLegend | HB-7 |
| Live/dead | UV Blue | N/A | Invitrogen (Thermo Fisher Scientific) | N/A |

**Supplementary Table 5.** Median proportions (%CD45+ cells) and P-values for significant cell types in nasal-bronchial, nasal-alveolar, bronchial-alveolar comparisons in Figure 5B.

| **Nasal-vs- bronchial** | | | |
|---|---|---|---|
| | **Median nasal** | **Median bronchial** | **FDR-P** |
| CD66blow CD16+ neutrophils | 12.56 | 3.60 | 0.02 |
| Macrophages | 0.99 | 3.46 | 0.01 |
| NK-T cells | 0.16 | 0.98 | 0.002 |
| **Nasal-vs-alveolar** | | | |
| | **Median nasal** | **Median alveolar** | **FDR-P** |
| B cells | 3.92 | 0.97 | 0.01 |
| Eosinophils | 0.22 | 0.07 | 0.01 |
| CD8 T cells | 20.71 | 3.87 | 0.0003 |
| Neutrophils | 28.17 | 1.82 | <0.000001 |
| CD66blowCD16+ neutrophils | 12.56 | 0.06 | <0.000001 |
| CD66bhighCD16+neutrophils | 4.49 | 0.85 | 0.007 |
| CD66bhighCD16- neutrophils | 1.77 | 0.11 | 0.00007 |
| CD66blowCD16- neutrophils | 2.17 | 0.06 | <0.000001 |
| Macrophages | 0.99 | 80.06 | <0.000001 |
| **Bronchial-vs-alveolar** | | | |
| | **Median bronchial** | **Median alveolar** | **FDR-P** |
| Neutrophils | 30.75 | 1.82 | 0.0002 |
| Eosinophils | 0.41 | 0.07 | 0.03 |
| CD66blowCD16+ neutrophils | 3.60 | 0.06 | 0.0005 |
| CD66bhighCD16+neutrophils | 6.86 | 0.85 | 0.04 |
| CD66bhighCD16- neutrophils | 8.00 | 0.11 | 0.002 |
| CD66blowCD16- neutrophils | 2.85 | 0.06 | 0.0002 |
| Macrophages | 3.46 | 80.06 | 0.00002 |
| NK cells | 1.46 | 0.39 | 0.01 |
| NK-T cells | 0.98 | 0.19 | 0.03 |
| CD8 T cells | 13.78 | 3.87 | 0.04 |

**Supplementary Table 6** Spearman coefficients and FDR-P values for the nasal-bronchial-blood immune cell axis in Figure 5C.

| Cell type | Spearman-ρ | FDR-P |
|---|---|---|
| **Nasal-bronchial** | | |
| Eosinophils | 0.85 | 0.002* |
| Neutrophils | 0.64 | 0.030* |
| Macrophages | 0.42 | 0.220 |
| NK cells | -0.18 | 0.610 |
| NK-T cells | 0.19 | 0.610 |
| CD8 T cells | 0.69 | 0.020* |
| CD4 T cells | 0.70 | 0.020* |
| Monocytes | 0.72 | 0.020* |
| B cells | 0.12 | 0.690 |
| cDCs | 0.92 | 0.0002* |
| **Nasal-blood** | | |
| Eosinophils | 0.44 | 0.16 |
| Neutrophils | -0.15 | 0.66 |
| Macrophages | NA | NA |
| NK cells | -0.20 | 0.65 |
| NK-T cells | -0.02 | 0.92 |
| CD8 T cells | -0.13 | 0.66 |
| CD4 T cells | -0.22 | 0.65 |
| Monocytes | 0.24 | 0.65 |
| B cells | -0.03 | 0.92 |
| cDCs | 0.13 | 0.66 |
| **Bronchial-blood** | | |
| Eosinophils | 0.35 | 0.59 |
| Neutrophils | 0.08 | 0.98 |
| Macrophages | NA | NA |
| NK cells | -0.20 | 0.85 |
| NK-T cells | 0.72 | 0.03* |
| CD8 T cells | -0.36 | 0.59 |
| CD4 T cells | -0.01 | 0.98 |
| Monocytes | -0.25 | 0.81 |
| B cells | 0.05 | 0.98 |
| cDCs | -0.07 | 0.98 |

**Supplementary Table 7.** Median proportions (%CD45+ cells) and P-values for significant cell types in tonsil-adenoid comparison in Figure 6B.

| | **Median tonsils** | **Median adenoids** | **FDR-P** |
|---|---|---|---|
| Neutrophils | 0.59 | 0.26 | 0.009 |
| Plasmablasts | 1.38 | 0.88 | 0.0008 |
| GC B cells | 18.84 | 10.38 | 0.002 |
| Classical memory B cells | 1.63 | 2.79 | 0.002 |
| Non-switched memory B cells | 1.69 | 2.72 | 0.002 |
| NK cells | 0.36 | 0.63 | 0.00005 |
| pDCs | 0.32 | 0.82 | 0.000001 |

**Supplementary Table 8.** Spearman coefficients and FDR-P values for tonsil-adenoid-blood immune cell axis in Figure 6C.

| | **Tonsil-adenoid** | | **Tonsil-blood** | | **Adenoid-blood** | |
|---|---|---|---|---|---|---|
| | Spearman-ρ | FDR-P | Spearman-ρ | FDR-P | Spearman--ρ | FDR-P |
| GC B cells | 0.28 | 0.166 | NA | NA | NA | NA |
| Plasma blasts | 0.60 | 0.002* | -0.26 | 0.288 | -0.03 | 0.892 |
| Atypical memory B cells | 0.80 | 0.000* | 0.54 | 0.046* | 0.55 | 0.013* |
| Classical memory B cells | 0.65 | 0.001* | 0.27 | 0.288 | 0.53 | 0.017* |
| Naive B cells | 0.59 | 0.002* | 0.37 | 0.185 | 0.31 | 0.201 |
| Non-switched memory B cells | 0.45 | 0.024* | 0.15 | 0.563 | 0.16 | 0.480 |
| Plasma cells | 0.51 | 0.010* | 0.28 | 0.288 | 0.46 | 0.040* |
| Transitional B cells | 0.55 | 0.005* | 0.61 | 0.016* | 0.37 | 0.109 |
| NK cells | 0.17 | 0.398 | 0.27 | 0.288 | 0.38 | 0.109 |
| NK-T cells | 0.64 | 0.002* | -0.05 | 0.830 | 0.30 | 0.201 |
| CD8 $T_{CM}$ | 0.62 | 0.002* | 0.44 | 0.089 | 0.48 | 0.038* |
| CD8 $T_{N}$ | 0.53 | 0.007* | 0.61 | 0.016* | 0.37 | 0.109 |
| CD8 $T_{EMRA}$ | 0.50 | 0.012* | 0.06 | 0.797 | 0.17 | 0.480 |
| CD8 $T_{EM}$ | 0.51 | 0.011* | 0.35 | 0.200 | 0.56 | 0.013* |
| Activated Treg | 0.55 | 0.005* | 0.44 | 0.089 | 0.30 | 0.201 |
| Naïve Treg | 0.68 | 0.001* | 0.50 | 0.070 | 0.72 | 0.000* |
| $T_{FH}$ | 0.61 | 0.002* | 0.48 | 0.071 | 0.27 | 0.245 |
| CD4 $T_{helper}$ | 0.34 | 0.098 | 0.26 | 0.288 | 0.16 | 0.480 |
| CD4 $T_{N}$ | 0.42 | 0.038* | 0.44 | 0.089 | 0.09 | 0.644 |
| CD4 $T_{EMRA}$ | 0.65 | 0.001* | 0.34 | 0.203 | 0.12 | 0.611 |
| Monocytes | 0.34 | 0.098 | -0.07 | 0.797 | 0.17 | 0.480 |
| cDCs | 0.21 | 0.313 | 0.27 | 0.288 | 0.10 | 0.640 |
| pDCs | 0.63 | 0.002* | 0.24 | 0.301 | 0.44 | 0.048* |
| ILCs | 0.60 | 0.002* | 0.08 | 0.797 | 0.20 | 0.444 |
| Neutrophils | 0.38 | 0.066 | 0.27 | 0.288 | -0.22 | 0.379 |

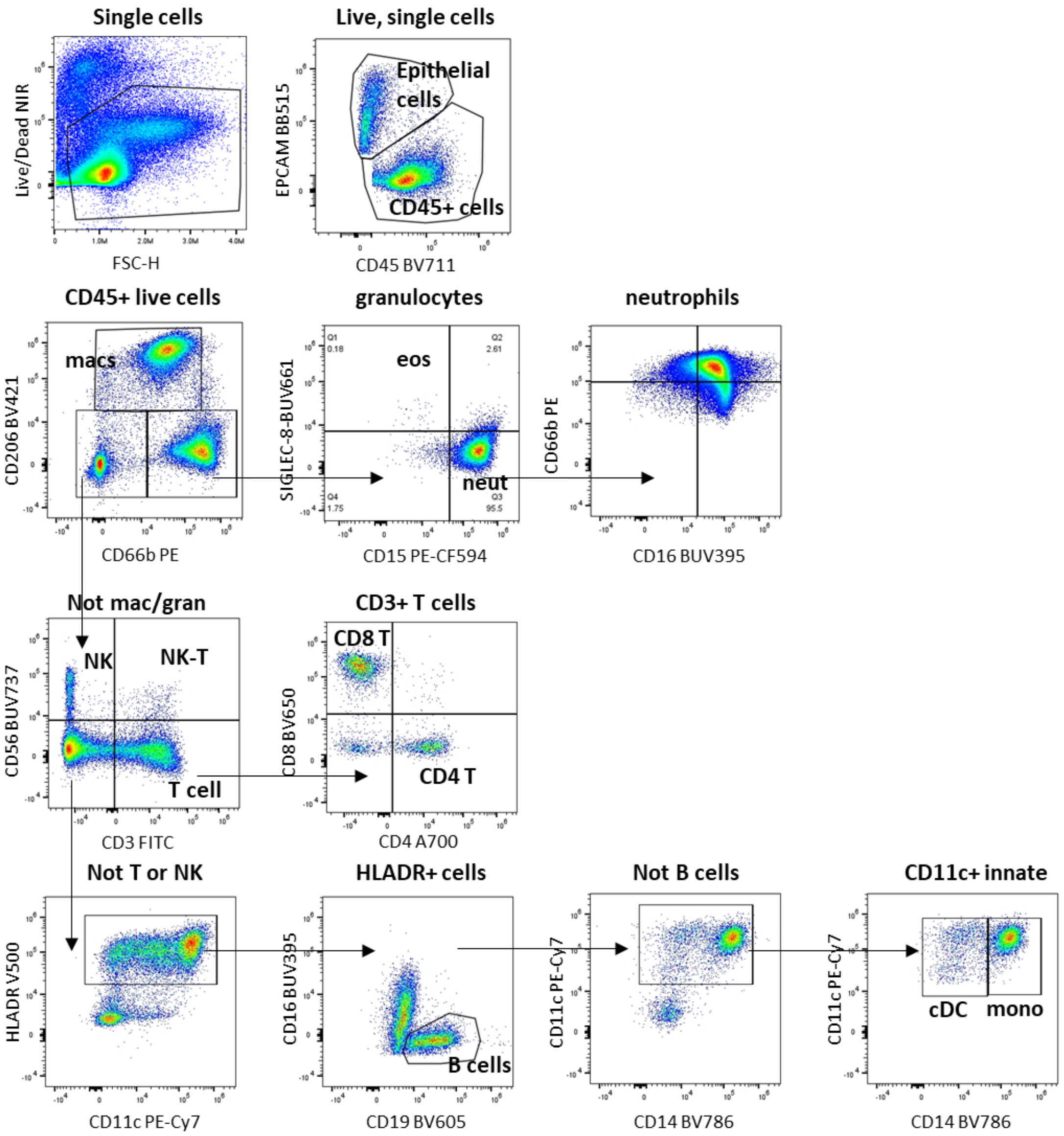


**Supplementary Figure 1.** Representative flow cytometry gating strategy for nasal, bronchial, and alveolar samples. Live cells were selected based on viability dye, following which expression of lineage markers CD45 and EPCAM were used to distinguish all immune cells (CD45+ cells) and all epithelial cells (EPCAM+ cells). Within CD45+ live cells, macrophages were selected based on CD206 expression, and total granulocytes were selected based on CD66b expression. Within the granulocyte population, eosinophils were selected based on SIGLEC-8 expression and neutrophils were selected based on CD15 expression. Neutrophil subsets were identified using CD66b and CD16. CD206- CD66b- cells were subtyped into CD56+ NK cells, CD56+CD3+ NK-T cells, and CD3+ T cells. Within the CD3+ T cell fraction, CD4 and CD8 T cells were identified. HLADR+CD19+ cells were identified as B cells. Monocytes were identified based on D14 expression from within the CD11c+ HLADR+ population. Finally, classical DCs (cDCs) were selected based on HLADR+CD14-CD11c+ expression pattern.

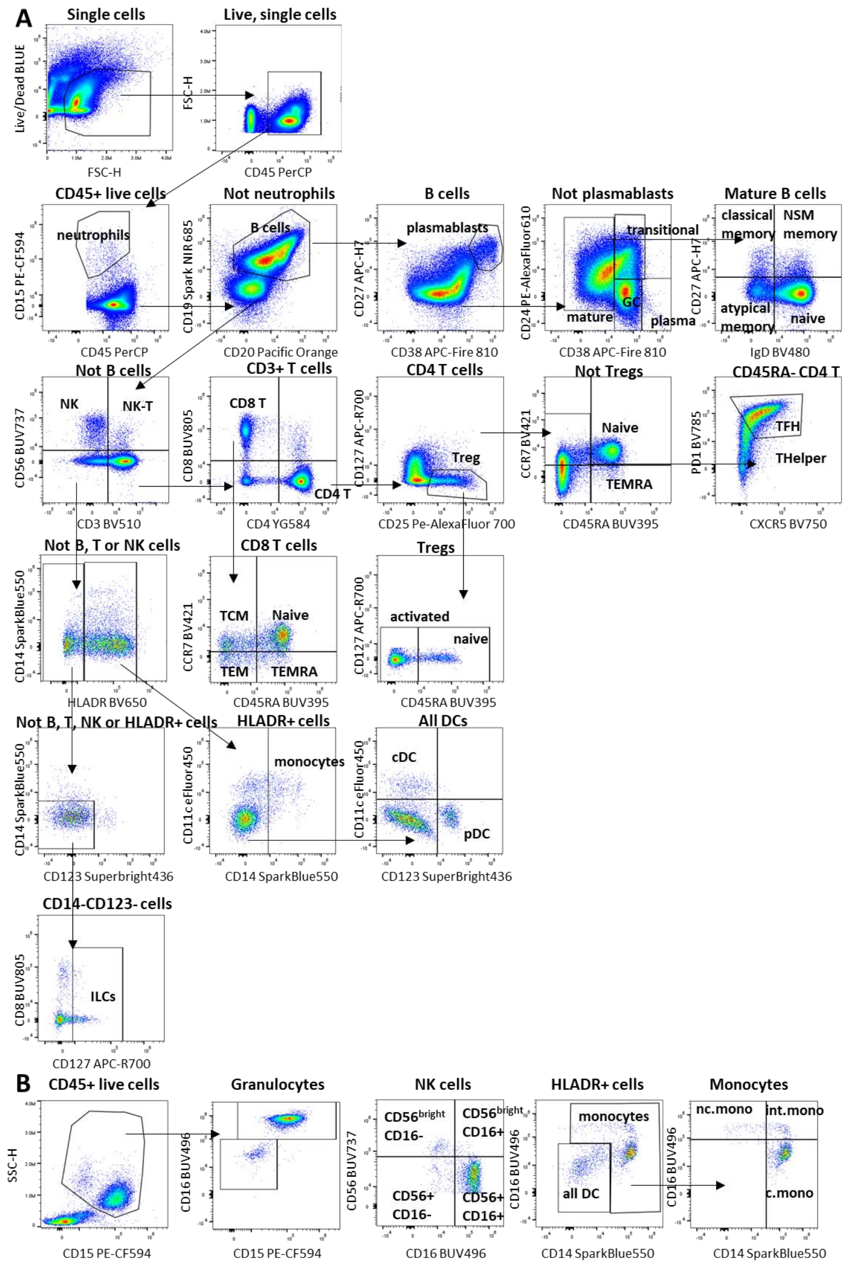


**Supplementary Figure 2.** Representative flow cytometry gating strategy for adenoid, tonsil, and whole blood samples. **A**. Gating strategies in common for all sample types: Live cells were selected based on viability dye, following which the lineage marker CD45 was used to distinguish all immune cells (CD45+ cells). Within the CD45+ immune population, neutrophils were identified based on CD15 expression. B cells were identified from the non-neutrophil population as CD19+ cells. Within the B cell population, plasmablasts ($CD38^{high}CD27^{high}$), mature (CD38-), germinal centre (CD38+CD24-), transitional (CD38+CD24+) and plasma cells ($CD38^{high}$, $CD24^{low}$) were identified. Within the mature B cell population, expression of IgD and CD27 was used to distinguish naïve, classical memory, non-switched memory and atypical memory B cells. Next, NK cells were identified from within the non-B cell fraction based on CD56 expression. NK-T cells expressed both CD3 and CD56. CD3-expressing T cells were identified as CD4 T cells or CD8 T cells. Within the CD4 T cell population, Tregs were identified based on a CD25+CD127- profile, and CD45RA was used to distinguish activated (CD45RA-) from naïve (CD45RA+) Tregs. Naïve and TEMRA CD4 T cells were identified from within the non-Treg CD4 fraction. CD45RA- CD4 T cells were classified as T-follicular helper (TFH) or Thelper based on expression of CXCR5 and PD1. CD8 T cells were classified as Naïve, TEMRA, TCM or TEM based on CCR7 and CD45RA expression. Next, CD19-CD20-CD3-CD56-HLADR+ cells were classified as monocytes (CD11c+CD14+), cDCs (CD11c+CD14-CD123-) or pDC (CD11c-CD14-CD123+). Finally, ILCs were distinguished as CD127+ cells that did not express any of the other lineage markers. **(B).** Gating strategies unique to whole blood samples: Both eosinophils (CD15lowCD16-) and neutrophils (CD15+CD16+) were identified from within a total granulocyte population. NK cells were separated into subsets based on expression of CD56 and CD16. These populations of NK cells were not evident in the tonsil or adenoid samples. Finally, three subsets of monocytes were identified based on CD14 and CD16 expression: classical monocytes, intermediate monocytes, non-classical monocytes. These populations of monocytes were not evident in the tonsil or adenoid samples.

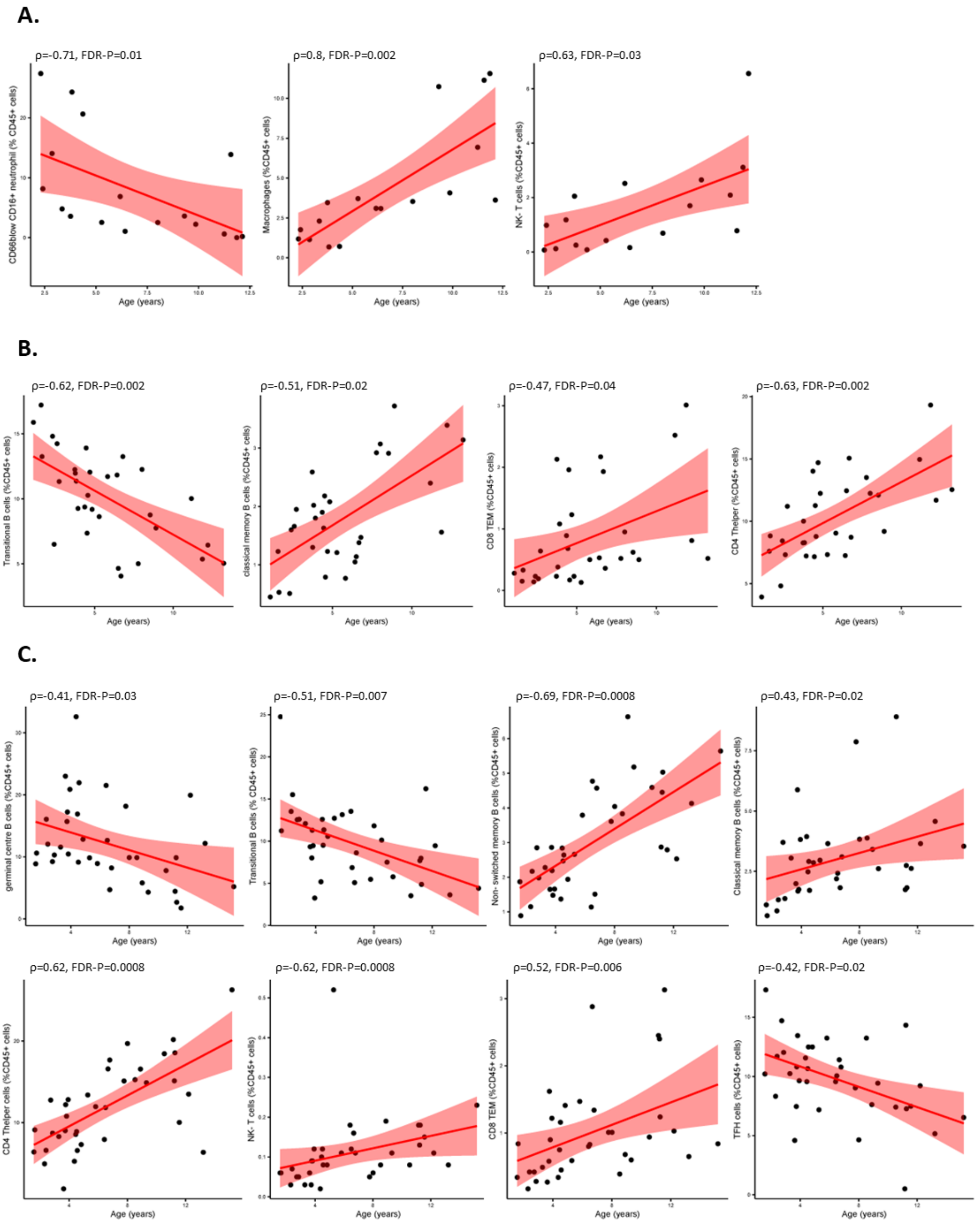


**Supplementary Figure 3.** Spearman correlation plots for each significant cell type in each tissue: **(A)** Bronchial brushings, **(B)** tonsils and **(C)** adenoids. The plots depict age in years on the x-axis, and the proportion of significant cell type on the y-axis. Spearman coefficient (ρ) and FDR-corrected p-values are shown.

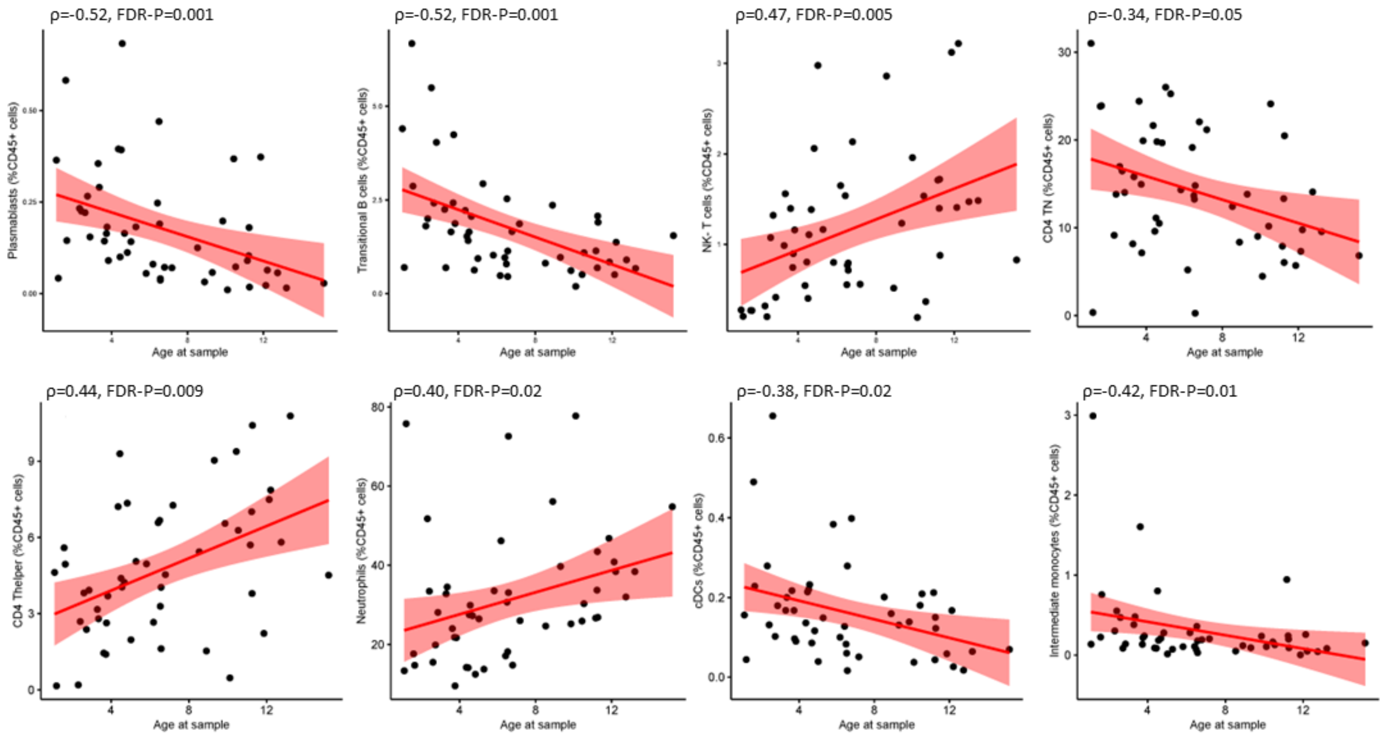


**Supplementary Figure 4.** Spearman correlation plots for each significant cell type in each whole blood. The plots depict age in years on the x-axis, and the proportion of significant cell type on the y-axis. Spearman coefficient (ρ) and FDR-corrected p-values are shown.

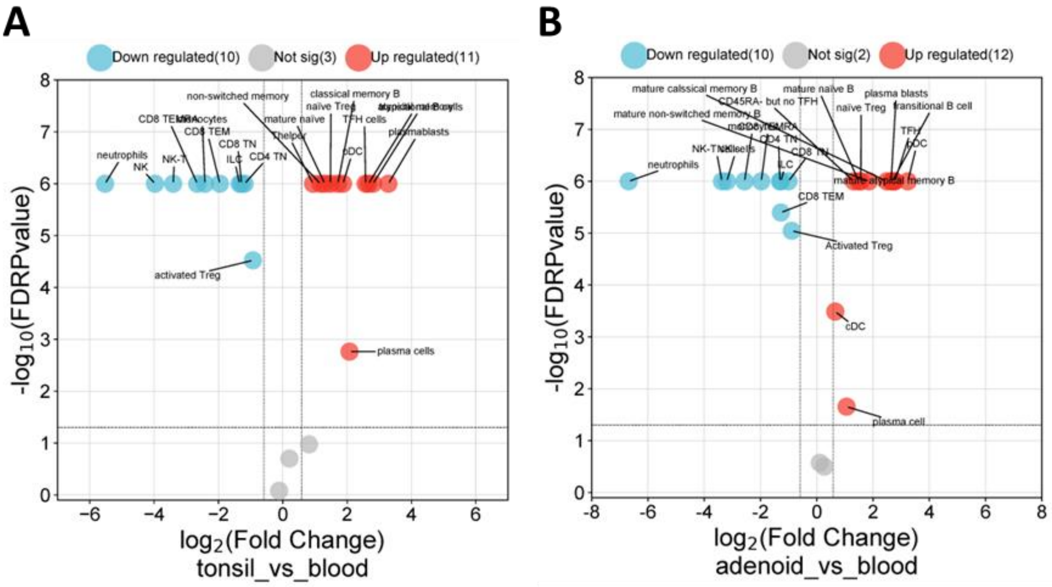


**Supplementary Figure 5.** Statistical comparison in immune cell proportions between **(A)** tonsils and blood, and **(B)** adenoids and blood. Cross-sectional p-values (part B) were determined by Mann-Whitney U-test and corrected for false discovery rate using the Benjamini Hochberg approach. FDR $p<0.05$ and fold change $>1.5$ was considered significant.
